# Gender-based violence rates and mental health disease burden in India, 2001-2021: A state-level ecological study

**DOI:** 10.64898/2026.10.16.26365723

**Authors:** Sakshi Sangekar, Gergo Baranyi, James B Kirkbride, Praveetha Patalay

## Abstract

Violence against women is a key contributor to mental health gender disparities, with greater gender gaps observed in low- and middle-income countries. This study examines state-level associations between gender-based violence against women and mental health burden in India over time. We linked 1) annual state-level age-standardised disability-adjusted life year rates (ASDR) from the Global Burden of Disease study for overall mental health disorders, depressive disorders, anxiety disorders, bipolar disorder, schizophrenia and self-harm, with 2) annual state-level gender-based violence rates from the Open Government Data Platform, India. We used descriptive ecological analyses and multilevel modelling to estimate the state-level association between gender-based violence and sex-stratified mental health outcomes over time (2001-2021).

Results showed higher state-level gender-based violence (GBV) rates were associated with higher state-level ASDR of schizophrenia (β_women_ = 0.39 [95%CI 0.25-0.52]; β_men_ = 0.42 [0.29-0.55]) and bipolar disorder (β_women_ = 0.02 [0.01-0.03]; β_men_ = 0.04 [0.03-0.05]), after adjustment for state-level social development, but not with other outcomes. Higher GBV rates were associated with a lower gender disparity in depressive disorders. Longitudinal findings revealed that the state-level association between GBV rates and ASDR of overall mental health disorders strengthened over time (β_women_ = 5.08 [3.96-6.20]; β_men_ = 3.38 [2.60-4.15]), whereas the association weakened for self-harm and schizophrenia. Lagged findings suggested that with an increase in GBV rate, ASDR of overall mental health disorders increased for men over two years and for both sexes over five years. Our findings highlight the importance of considering state-level trends in GBV and related policies, and considering impacts of GBV on both men and women’s mental health.

## Introduction

Globally, women are twice as likely as men to suffer from common mental health conditions, with a wider gender gap observed in low-and middle-income countries.^1,2^ In India, the overall mental health burden has doubled from 2.5% in 1990 to 4.7% in 2017, with sub-national variations in mental health gender disparities.^3,4^ Recent research increasingly recognises gender-based violence as a key contributor to worsening mental health among Indian women compared with men.^2,5^

In the Indian context, gender-based violence (GBV) remains a complex public health issue, rooted in longstanding socio-cultural practices such as child marriage, honour killings, dowry-related practices, female infanticide, and female genital mutilation.^6–8^ Evidence highlights that dowry-related humiliation, sexual violence, intimate partner violence, domestic abuse, and economic abuse perpetrated by husbands are linked to anxiety, depressive disorders, and suicidal ideation, including attempted suicide, among Indian ever-married women.^9–12^

Despite the Indian government introducing initiatives such as the One Stop Centre Scheme 2015, the Mahila Police Volunteer Scheme 2017, and the Integrated Women Empowerment Programme 2020, for women’s safety and emergency aid, violence against women remains persistently high, with state-level differences in the implementation of these schemes.^13,14^ Additionally, state-level differences in institutional factors, such as the availability of reporting infrastructure and law enforcement, highlight the importance of assessing region-specific changes in the reported incidences of gender-based violence and their impact on mental health burden.^13,15,16^

Existing GBD studies in India have revealed that the distribution of mental health disorders is substantially influenced by state-level socio-demographic factors, such as fertility rates, literacy rates, and income per capita.^3,17^ For instance, southern states with better socio-demographic development report higher prevalence of mental health disorders, including depressive and anxiety disorders, compared with northern states with worse socio-demographic development.^3,17^

Additionally, researchers have observed an increase in mental health burden, including self-harm, linked with increases in reported cases of domestic abuse and violence.^18,19^ It is therefore crucial to understand the distribution of mental health disease burden through an eco-social perspective, which suggests that mental ill-health is an interplay of structural, social and ecological stressors.^20^ Considering India’s state-level differences in governance, urbanisation, healthcare and safety infrastructure, it is necessary to examine the cumulative effect of these stressors on the distribution of mental health disease burden at state-level across time points.

Although existing studies have examined the mental health consequences of various forms of abuse, they primarily focus on domestic and intimate partner violence reported by ever-married women.^6,10,21^ There is a broader need to examine the systemic nature of violence against women by including population estimates of violence against women (e.g. sex trafficking, acid attacks, and dowry deaths) and assessing its region-specific impact on the mental health burden over time. To address this research gap, we linked state-level crime records of violence against women in India with estimates of mental health disorders.

We aimed to answer four key research questions within this context: (a) Is there an association between state-level GBV against women and the state-level burden of mental health disorders among women and men in India? (b) Is there an association between state-level GBV against women and gender disparities in the state-level burden of mental health disorders? (c) Has the association between state-level GBV against women and the state-level burden of mental health disorders among women and men changed over time in India? (d) Is there a temporal association between state-level gender-based violence against women and the state-level burden of mental health disorders among men and women in India from 2001 to 2021?

## Methods

### Data sources

We obtained state-level estimates of gender-based violence against women from the National Crime Records Bureau (NCRB), available on the Open Government Data (OGD) Platform India (https://www.data.gov.in/). As gender identity data were not readily available in national and state-level datasets, we used state-level records of crime against women from 2001 to 2021 to calculate annual rates of gender-based violence. We retrieved age-standardised disability-adjusted life-year rates (ASDR) to measure the state-level burden of mental health disorders from the GBD Study (https://ghdx.healthdata.org/), maintained by the Institute for Health Metrics and Evaluation (IHME) at the University of Washington. This study adhered to the Guidelines for Accurate and Transparent Health Estimates Reporting (GATHER) Standards to ensure transparency and accuracy in reporting population-level data across time points and geographical units (supplement pp 25).^22^

### Procedure

We included a total of 30 geographical units spanning India: 28 States (Andhra Pradesh, Telangana, Arunachal Pradesh, Assam, Bihar, Chhattisgarh, Goa, Gujarat, Haryana, Himachal Pradesh, Jharkhand, Karnataka, Kerala, Madhya Pradesh, Maharashtra, Manipur, Meghalaya, Mizoram, Nagaland, Odisha, Punjab, Rajasthan, Sikkim, Tamil Nadu, Tripura, Uttar Pradesh, Uttarakhand, West Bengal) and two Union Territories (Jammu & Kashmir and Delhi). Union Territories (UTs) are smaller geographical units than Indian states and are governed by the Central Government of India. The remaining five UTs were excluded due to the unavailability of disaggregated data.

### Exposure: State-level gender-based violence (GBV) rate

According to NCRB, crimes against women include rape, kidnapping and abduction, dowry-related crimes (including dowry deaths), molestation, sexual harassment, assault on women with intent to outrage her modesty, insult to the modesty of women, cruelty by husband or his relatives (domestic violence), importation of girls, immoral trafficking, indecent representation of women, and sati practices (refers to a widow burning herself on her husband’s funeral pyre).^23^

As mid-year population was unavailable in the NCRB data for the years 2012, 2013 and 2014, we used mid-year population estimates from the National Health Profiles 2013 and 2018.^24,25^ GBV estimates for Andhra Pradesh and Telangana differed due to state reorganisation; the method is shown in the supplement (pp. 3). The following formula was used to estimate the annual state-level GBV rate:

*State-level GBV rate = (annual state-level reported cases of total crimes against women / State-specific mid-year population) x 100,000*

### Outcomes: State-level burden of mental health disorders and gender disparities

We used age-standardised disability-adjusted life-years rates (ASDR) to measure the burden of six mental health outcomes among women and men across the 30 geographical units: overall mental health disorders, schizophrenia, bipolar disorder, depressive disorders, anxiety disorder and self-harm. GBD calculated ASDR as the sum of years lost due to premature death and years lived with disability, per 100,000 population (supplement pp 2). GBD data categorised mental health disorders using the diagnostic criteria of DSM-IV-TR (Diagnostic and Statistical Manual of Mental Disorders, Fourth Edition) and ICD-10 (International Classification of Diseases and Related Health Problems, 10th Revision) (supplement pp. 3).

To quantify state-level gender disparity, we calculated the gender disparity rate ratio (GDR) for each of the six mental health outcomes using the following formula:

*State-level GDR = annual state-level ASDR for women /annual state-level ASDR for men*

A GDR of 1 represented an equal burden between men and women, a GDR greater than 1 represented a higher burden among women, and a GDR less than 1 represented a higher burden among men.

### Covariate: State-level socio-demographic Index (SDI)

We used SDI from the GBD study, which provides a composite indicator of socio-demographic development status for each state. SDI is the geometric mean of (a) total fertility rate under age 25, (b) mean education for those aged 15 and older, and (b) lag-distributed income per capita. SDI values range from 0, indicating the lowest level of socio-demographic development, to 1, indicating the highest level of socio-demographic development.^26^ SDI estimates were unavailable for the year 2020, and we used linear interpolation to estimate these.

### Statistical Analysis

First, we calculated the state-level means with 95% confidence intervals (CIs) for aggregate GBV rates, overall mental health disorders ASDR, and gender disparity rate ratio from 2001 to 2021. The same was reported for the remaining five mental health outcomes provided in the supplement (pp 4-8).

Second, we used scatterplots to examine the relationship between GBV rates and each of the six mental health outcomes for women, men and the gender disparity. We present state-specific maps of India and visualise changes in GBV rates and gender disparities across all mental health outcomes at two time points: 2001 and 2021. Third, we used line graphs to visualise changes in state-level trends in GBV rates and each of the six mental health outcomes from 2001 to 2021.

We used a multilevel modelling approach to account for the hierarchical structure of the state-level data, i.e., 30 geographical units over time (2001-2021). GBV rates were rescaled per 10,000 population to aid the interpretability of effect size estimates. In a null model, we estimated intra-class correlation coefficients (ICC) for the proportion of total variance in each mental health outcome explained by state-level clustering, as provided in the supplement (pp 9). ICC values ranged from 0 (lowest) to 1, with higher values indicating a greater proportion of the variability in rates was attributable to between-state differences.^27^ Next, we fitted linear mixed effects analyses to investigate state-level associations between GBV rates and mental health outcomes among women, men and the gender disparity rate ratio. Year and State-level SDI were included in the mixed-effects models as fixed effects. To examine whether the association between GBV rates and six mental health outcomes changed over time, we included an interaction between GBV and year in the above models, with year centered on 2001.

To examine temporal associations, we conducted SDI-adjusted fixed-effects analyses, applying a 2-year time lag (i.e., t-2) model to the exposure-GBV rate. The fixed-effects models were repeated using t-1 and t-5 to assess time sensitivity (see supplement pp 10; table S7). We also reran the models after excluding data from 2020 and 2021, which may have been affected by COVID-19 pandemic-related sensitivities (supplement pp 11; table S8). All analyses were performed in Stata version 18.5.

## Results

### Descriptives

GBV rates varied considerably across Indian states, ranging from 46.5 GBV cases per 100,000 population in Assam to 11.1 GBV cases per 100,000 population in Bihar (Table 1). Scatterplots in Figure 1 showed no evidence of a correlation between GBV rate and overall mental health disorder ASDRs for both women (r= -0.001, p= 0.99) and men (r= -0.012, p= 0.95).

**Table 1.**
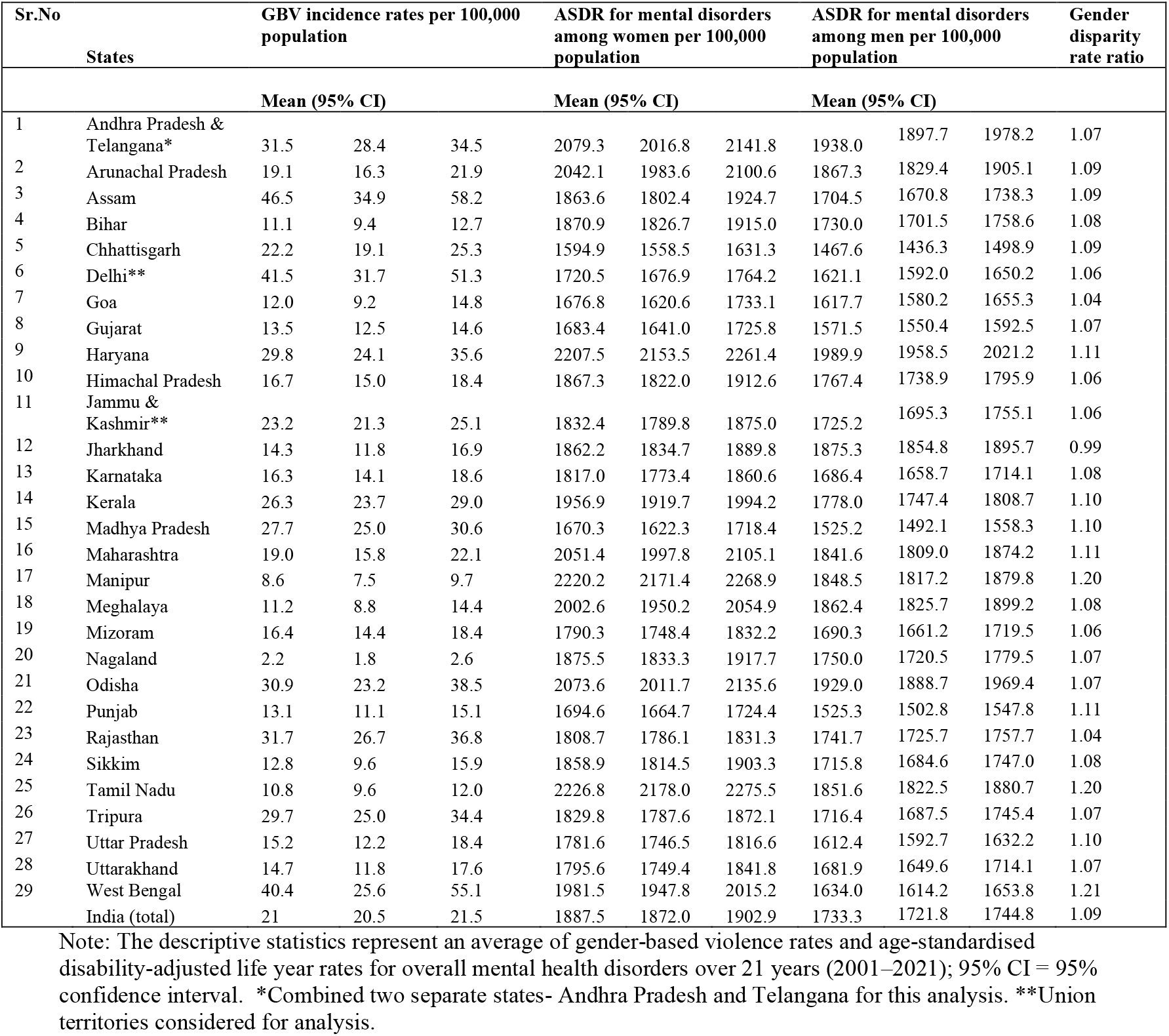
Descriptive statistics representing the average of gender-based violence rates, age-standardised disability-adjusted life years for overall mental health disorder burden among women and men per 100,000 population, and the gender disparity rate ratio by 30 geographical units (Indian States/Union Territories), 2001-2021.

**Figure 1.**
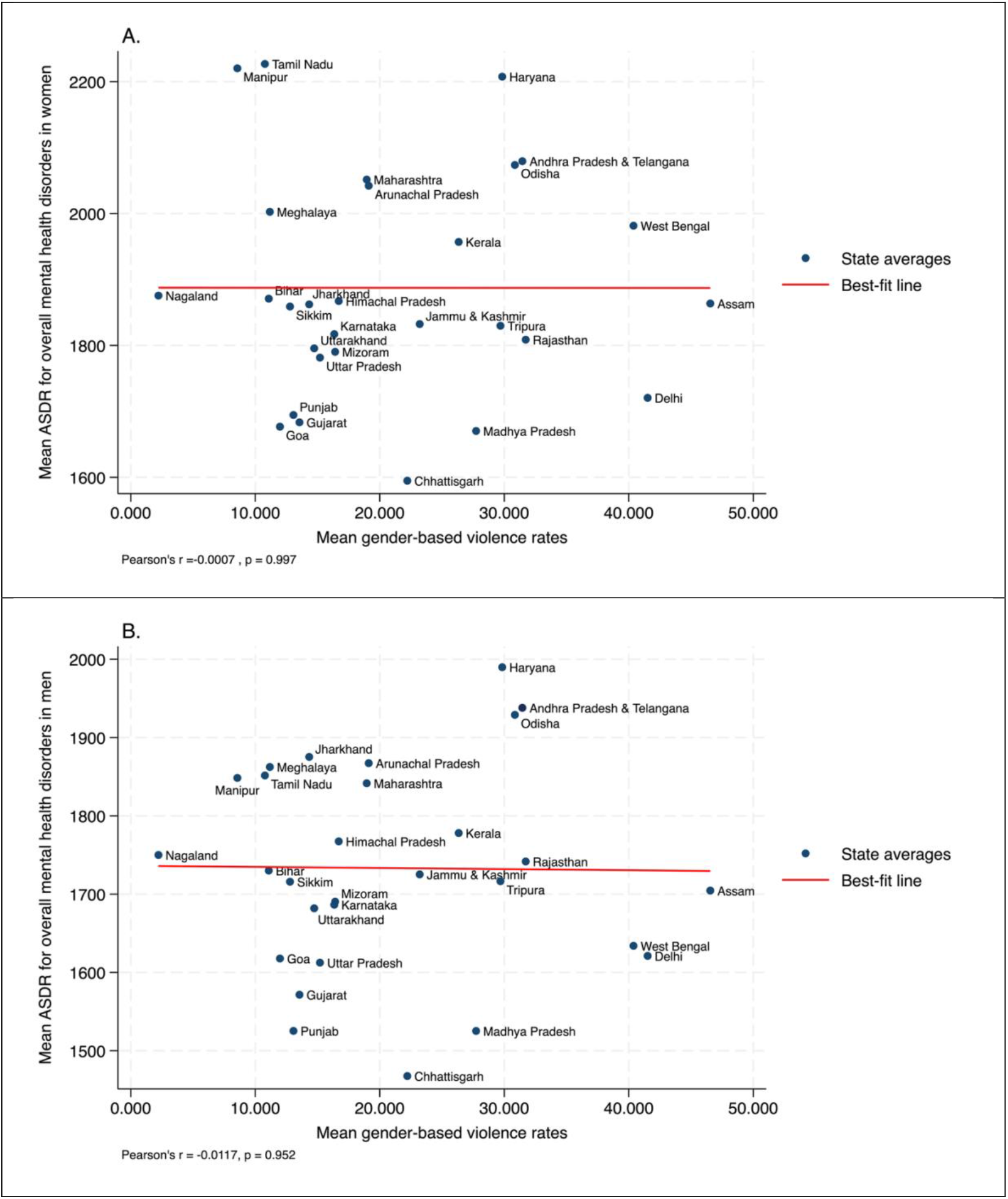
Scatterplot showing the relationship between mean gender-based violence rates and mean age-standardised disability rates for overall mental health disorders in women (A) and men (B) over time (2001-2021) for 30 geographical units (Indian states/Union Territories). Disorder-specific plots are available in the Supplement (Figures S1.1 – S1.3) Note. Each dot represents one of the 30 geographical units (states/Union territories), with values shown as cumulative means over 21 years. The x-axis denotes mean gender-based violence rates, and the y-axis denotes mean age-standarised disability-adjusted life year rates (ASDR) for overall mental health disorders among women (A) and men (B). The red line represents the line of best fit, illustrating the trend and strength of correlation between mean gender-based violence rates and ASDR. In women (A), the correlation coefficient was Pearson’s r = –0.0007 (p = 0.997), and in men (B), Pearson’s r = –0.0117 (p = 0.952), indicating no statistically significant association in either group.

Figure 2 showed higher GBV cases across Indian states in 2021 compared to 2001, with substantial increases in GBV rates in West Bengal, Odisha, and Assam over time. In 2021, West Bengal indicated the highest gender disparity in overall mental health disorders (1.25), depressive disorders (1.62) and anxiety disorders (1.74), wherein for every 100 male, 162 women would be affected by the burden of depressive disorders and 174 women affected by the burden of anxiety disorders (figure 2 and supplement pp 15-16; figures S2.1 and S2.2), and this disparity remains relatively unchanged between 2001 and 2021. Manipur (1.26) and Tamil Nadu (1.22), despite lower GBV rates, indicated relatively higher gender disparities in ASDR for overall mental health disorders (Figure 2).

**Figure 2.**
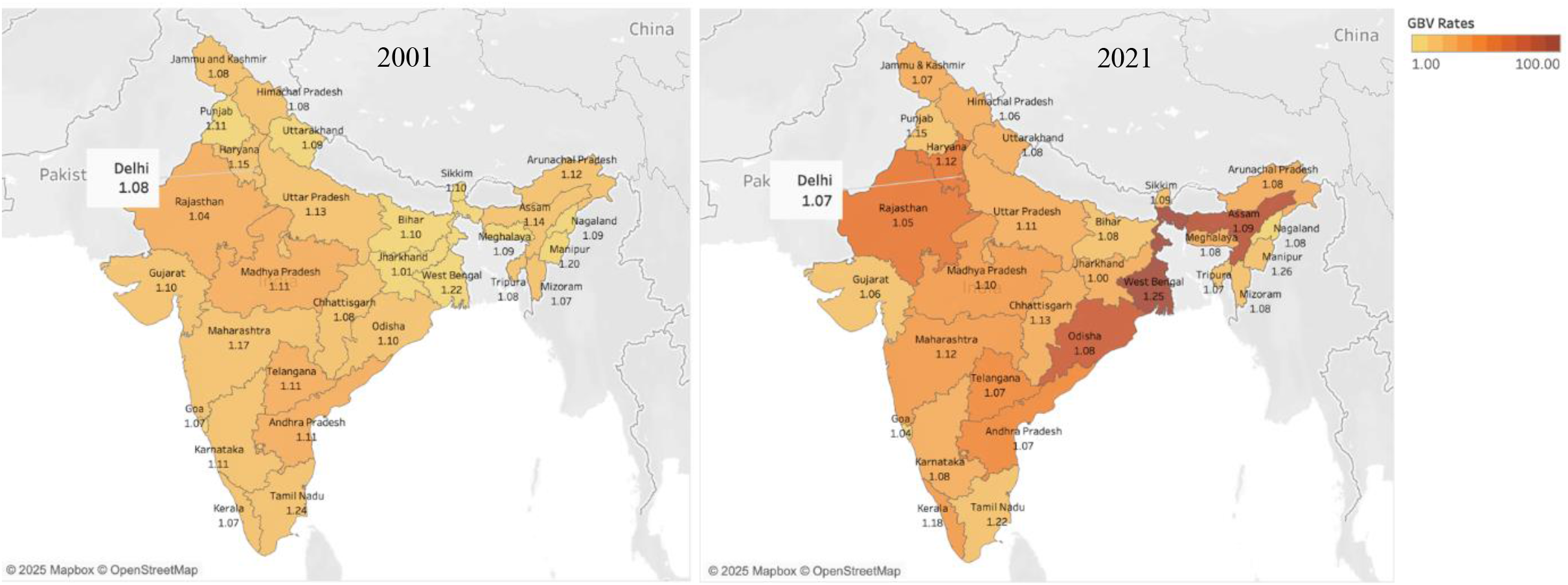
Distribution of gender-based violence rates and gender disparity in overall mental health disorder burden across 30 geographical units (Indian states/ Union Territories) at two time points, 2001 and 2021.Disorder-specific maps are available in the Supplement (Figures S2.1 – S2.5) Note: The Indian map displays gender-based violence rates by colour gradient, with darker shades denoting higher rates and lighter shades denoting lower rates. Each geographical unit is labelled with the gender disparity rate ratio of overall mental health disorder burden at baseline (2001) and the most recent year (2021). A gender disparity rate ratio greater than 1 denotes a higher mental health burden among women than men per 100,000 population.

Figure 3 showed considerable heterogeneity in the distribution of ASDR for overall mental health disorders across 30 geographical units from 2001 to 2021. Andhra Pradesh and Telangana, Arunachal Pradesh, Assam, and Meghalaya reported a significant decrease in the overall mental health burden between 2006 and 2009. In contrast, Manipur, Punjab, and West Bengal exhibited minimal changes in overall mental health burden during the same period (Figure 3). Assam, Meghalaya, Mizoram, Kerala, Manipur, and Nagaland continued to observe an upward trend in the burden until 2021. Conversely, the burden of overall mental health disorders decreased and plateaued in Tripura, Uttar Pradesh, Jharkhand, and Sikkim. West Bengal, Jammu and Kashmir, and Gujarat recorded a decline in 2021 (Figure 3).

**Figure 3.**
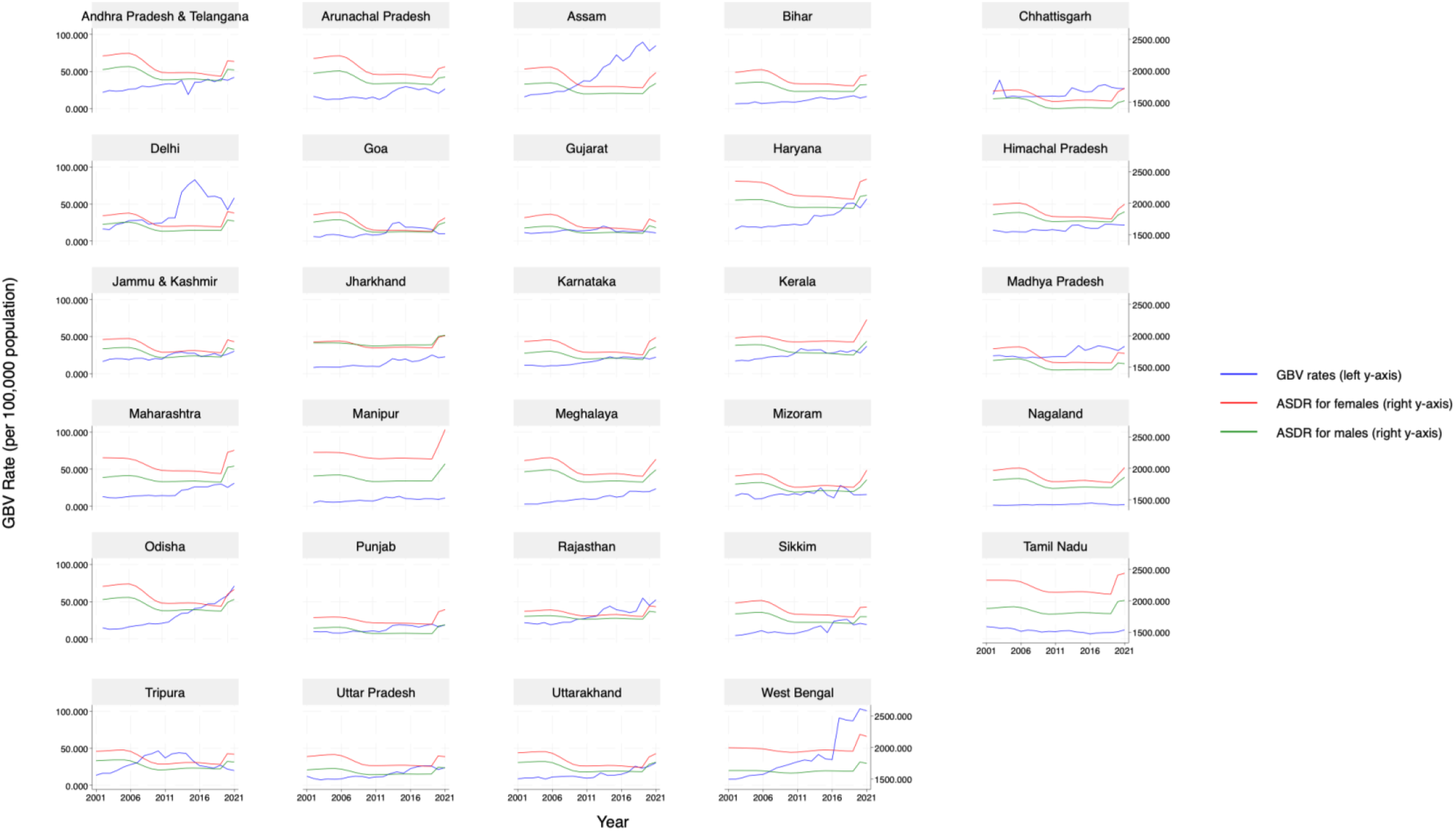
Line graphs showing trends in gender-based violence rates and age-standardised disability-adjusted life year rates of overall mental health disorders across geographical units (Indian states/Union Territories) from 2001 to 2021. Note: Each panel shows one of the 30 geographical units (Indian states/Union territories). Andhra Pradesh and Telangana are combined in one panel. The x-axis denotes the year from 2001 to 2021. The left y-axis represents gender-based violence (GBV) rates per 100,000 population, plotted as blue lines. The right y-axis represents age-standardised disability-adjusted life year rates (ASDR) of overall mental health disorders for women/females (red lines) and men/males (green lines) per 100,000 population. Each panel illustrates state-specific changes in GBV rates and overall mental health disorder ASDRs over time from 2001 to 2021.

### State-level findings

We found evidence of associations between state-level GBV rates and ASDR for schizophrenia and bipolar disorders, for both men and women, in unadjusted and adjusted models (Table 2). For example, in adjusted models, for every 1-unit increase in state-level GBV rate, state-level ASDR for schizophrenia among women increased by 0.38 units (95%CI 0.25-0.52) and among men increased by 0.42 units (0.29-0.55) per 100,000 population. For every 1-unit increase in state-level GBV rate per 10,000 population, state-level ASDR for bipolar disorder among women increased by 0.02 units (0.01-0.03) and among men increased by 0.039 units (0.03-0.05) per 100,000 population. We found no evidence of association between GBV rates and ASDR for all mental health disorders, or separately for depressive disorders, anxiety disorders or self-harm. For every 1-unit increase in state-level GBV rate per 10,000 population, state-level ASDR for bipolar disorder among women increased by 0.02 units (0.01-0.03) and among men increased by 0.04 units (0.03-0.05) per 100,000 population.

**Table 2.** Predicting six mental health outcomes from gender-based violence rates per 10,000 population among women and men from 2001 to 2021.

| Outcomes | Mixed effects unadjusted model |  |  | Mixed effects adjusted model for SDI |  |  | Fixed effects model (lag t-2) |  |  |
| --- | --- | --- | --- | --- | --- | --- | --- | --- | --- |
|  | women | men | Gender disparity rate ratio | women | men | Gender disparity rate ratio | women | men | Gender disparity rate ratio |
| GBV rate as exposure, Estimate; 95% CI=LL, UL |  |  |  |  |  |  |  |  |  |
| Mental health disorders | 3.00<br>(-5.26, 11.26) | 3.47<br>(-2.17, 9.11) | -0.001<br>(-0.002, 0.001) | 2.93<br>(-5.23, 11.10) | 3.44<br>(-2.16, 9.05) | -0.001<br>(-0.002, 0.001) | 15.82<br>(-2.73, 34.37) | 11.84*<br>(0.52, 23.169) | 0.001<br>(-0.003, 0.005) |
| Depressive disorders | 2.03<br>(-4.78, 8.84) | 2.43<br>(-2.53, 7.39) | -0.004**<br>(-0.007, -0.001) | 2.00<br>(-4.78, 8.79) | 2.42<br>(-2.53, 7.38) | -0.004**<br>(-0.007, -0.001) | 11.00<br>(-4.44, 26.44) | 8.66<br>(-1.85, 19.18) | -0.003<br>(-0.009, 0.003) |
| Anxiety disorders | -0.32<br>(-2.52, 1.88) | -0.37<br>(-1.63, 0.89) | 0.001<br>(-0.002, 0.004) | -0.33<br>(-2.47, 1.81) | -0.38<br>(-1.62, 0.87) | 0.001<br>(-0.002, 0.004) | 3.09<br>(-0.43, 6.62) | 1.57*<br>(0.17, 2.96) | 0.003<br>(-0.005, 0.011) |
| Self-harm | -5.87<br>(-14.51, 2.76) | -4.16<br>(-11.26, 2.94) | -0.002<br>(-0.009, 0.006) | -5.88<br>(-14.47, 2.72) | -4.15<br>(-11.22, 2.91) | -0.002<br>(-0.009, 0.006) | -4.63<br>(-36.77, 27.52) | -8.99<br>(-26.70, 8.71) | -0.002<br>(-0.019, 0.014) |
| Bipolar disorder | 0.02***<br>(0.01, 0.03) | 0.04***<br>(0.03, 0.05) | -0.000<br>(-0.000, 0.000) | 0.02***<br>(0.01, 0.03) | 0.04***<br>(0.03, 0.05) | -0.000<br>(-0.000, 0.000) | 0.02<br>(-0.02, 0.05) | 0.03<br>(-0.00, 0.06) | -0.000<br>(-0.001, 0.000) |
| Schizophrenia | 0.38***<br>(0.24, 0.53) | 0.42***<br>(0.28, 0.56) | 0.0003*<br>(0.0001, 0.0006) | 0.39***<br>(0.25, 0.52) | 0.42***<br>(0.29, 0.55) | 0.0003**<br>(0.0001, 0.0006) | 0.35<br>(-0.02, 0.71) | 0.37<br>(-0.03, 0.76) | 0.000<br>(-0.000, 0.001) |

Note: estimate=coefficient, \*p<0.05; \*\*p<0.01, \*\*\*p<0.001; 95% CI= 95% confidence interval; LL=lower limit, UL=upper limit, GBV rates= gender-based violence incidence rates, SDI=socio-demographic index values. Note: GBV rate rescaled from 100,000 population to 10,000 population.

In terms of gender disparities, depressive disorders ASDR was lowered by -0.004 per 100,000 population (-0.007 to -0.001) for every 1-unit increase in state-level GBV rate per 10,000 population (Table 2). Conversely, the state-level gender disparity for schizophrenia ASDR was increased slightly by 0.0003 (0.0001-0.0006) per 100,000 population for every 1-unit increase in state-level GBV rate per 10,000 population. No other associations were observed.

### Longitudinal findings

GBV rate and year interactions showed that the association between state-level GBV rates and ASDR for mental health disorders changed over time (Table 3). The state-level association between GBV rate and ASDR for mental health disorders strengthened by 5.08 units (3.96-6.20) for women and by 3.38 units for men (2.60-4.15) per year from 2001 to 2021. Similarly, the association between state-level GBV and the state-level burden of depressive disorders (β_women_ = 4.08 [3.14-5.02]; β_men_ = 2.87 [ 2.18-3.55]) and anxiety disorders (β_women_ = 0.90 [0.59-1.20]; β_men_ = 0.37 [0.19-0.55]) strengthened with every consecutive year from 2001 to 2021 (Table 3). Whereas the association of state-level GBV rate with the state-level burden of self-harm (β_women_= -2.69 [-3.92; -1.45]; β_men_= -2.22 [-3.24; -1.21]) and schizophrenia (β_women_= -0.02 [-0.04; -0.00]; β_men_= -0.02 [-0.04; -0.00]) weakened each year (Table 3).

**Table 3.** Interaction effects of state-level gender-based violence rates and year on state-level mental health outcomes, estimated using sex-specific mixed-effects adjusted models.

| Mental health outcomes | Interaction effect of GBV rate by Year (95%CI) |  |
| --- | --- | --- |
|  | women | men |
| Mental health disorders | 5.08*** (3.96-6.20) | 3.38*** (2.60-4.15) |
| Depressive disorders | 4.08*** (3.14-5.02) | 2.87*** (2.18-3.55) |
| Anxiety disorders | 0.90*** (0.59-1.20) | 0.37*** (0.19-0.55) |
| Self-harm | -2.69*** (-3.92; -1.45) | -2.22*** (-3.24; -1.21) |
| Bipolar disorder | -0.00 (-0.00; -0.00) | -0.00 (-0.00; -0.00) |
| Schizophrenia | -0.02* (-0.04; -0.00) | -0.02* (-0.04; -0.00) |

Note: estimate=coefficient, \* $p_{\text{interaction}} < 0.05$ ; \*\* $p_{\text{interaction}} < 0.01$ , \*\*\* $p_{\text{interaction}} < 0.001$ ; 95% CI= 95% confidence interval; GBV rates= gender-based violence incidence rates. The mixed-effects model is adjusted for state-level socio-demographic index (SDI). Note: GBV rate rescaled from 100,000 population to 10,000 population. The year is centred on 2001 as the baseline.

Fixed-effects analyses with a two-year lag showed that for every 1-unit increase in GBV rate per 10,000 population, the ASDR for mental health disorders increased by 11.84 units (0.52-23.17) and the ASDR for anxiety disorder increased by 1.56 units (0.17-2.96) among men over two years (Table 2). However, findings showed no evidence of an association among women.

### Sensitivity Analyses

Sensitivity analyses showed that an increase in state-level GBV rates was associated with an increase in the ASDR for mental health disorders among women (β = 19.19 [4.02-34.36]) and men (β = 14.49 [3.92-25.06]) over five years (Supplement pp 10; Table S7). Disorder-specific patterns were observed: higher GBV rates were associated with increased ASDR for schizophrenia among women (β = 0.44 [0.05-0.82]) and men (β = 0.46 [0.05-0.87]) over one year, whereas depressive disorders showed delayed effects, with elevated ASDR for women (β = 14.45 [0.78-28.13]) and men (β = 11.23 [1.09-21.38]) over five years (Supplement pp 10; Table S7).

After we dropped the observations from the COVID-19 pandemic, i.e., the years 2020 and 2021, results showed a positive state-level association between GBV rates and ASDR for mental health disorders among men (β = 5.02 [1.10-8.95]) (Supplement pp 10; Table S8). Compared with the main findings in Table 1, ASDR for schizophrenia showed larger effect sizes for both sexes (β_women_ = 0.52 [0.36-0.67]) (β_men_ = 0.56 [0.41-0.71]) (Supplement pp 11; Table S8).

## Discussion

In this ecological study using nationwide Global Burden of Disease data from India, we found that higher rates of state-level GBV were strongly associated with higher age-standardised disability rates for schizophrenia, and to a lesser extent bipolar disorder, for both men and women separately. We found that GBV rates had generally increased in India between 2001 and 2021, and that state-level associations between GBV and mental health outcomes had generally strengthened over this period. Finally, lagged analyses suggested that GBV rates both two and five years prior to the year in which ASDRs for mental health disorders were estimated revealed stronger associations between GBV and several mental health outcomes than cross-sectional comparisons.

In terms of gender disparity, we found higher GBV rates were linked to higher gender disparity in the burden of schizophrenia but lower gender disparity in the burden of depressive disorders at state-level. Lower gender disparity in depressive disorders may not necessarily indicate improved female outcomes but rather elevated male burden in states with high GBV rates. Prior research suggested that the presence of socio-economic stressors, such as exposure to violence, tends to increase the odds of depression among men and their likelihood of perpetrating violence.^5,28^ Another potential explanation could be that the high mortality risk of schizophrenia is more likely to be captured by ASDRs and reflected in the state-level data as premature deaths. Whereas the burden for anxiety and depressive disorders could be better captured by years lived with disability (YLDs) estimates.^29^

Previous studies have shown a strong association between gender-based violence against women and an increased risk of common mental health conditions such as depression and anxiety among women.^3,12,21^ Although our findings showed no evidence to support this concurrent relationship at the state level, GBV rates were observed to be strongly associated with ASDRs of mental health disorders over time for both sexes. For example, increases in state-level GBV rates were associated with a substantial increase in state-level ASDR for mental health disorders among women five years later; this may suggest that individual-level survey studies often capture women currently experiencing mental health issues following a history of violence, while ecological studies reveal a delayed impact rather than a concurrent effect. It is therefore important to interpret our findings cautiously.

Our study has various notable strengths. First, the use of longitudinal data spanning two decades (2001-2021) provided a more granular understanding of state-specific temporal variations in the association of GBV rates and mental health outcomes. Second, leveraging the National Crime Records provided a novel approach to capture the broader, multidimensional construct of gender-based violence, in contrast to previous studies that relied primarily on intimate partner violence data from the NFHS.^15,16^ Utilising data from all Indian States also enabled us to capture regional diversity, thereby increasing the relevance of our findings for policymakers and for designing targeted psychosocial interventions. Third, we controlled for socio-demographic index (SDI) at state-level and longitudinally over 20 years. Previous evidence shows that more developed states often report higher mental health burden due to better healthcare infrastructure; thus, controlling for SDI helped mitigate confounding bias and strengthened internal validity.^3,17^ Moreover, using ASDR from GBD accounted for state-level differences in population age structures, allowing for better comparison of disorder-specific temporal trends across the 30 geographical units.

Study limitations include use of state-level measures, which raises the risk of ecological fallacy, as aggregate state-level associations cannot be directly inferred to individuals.^30^ Nevertheless, regional-level comparisons over time are useful to elucidate state-specific contextual associations between gender-based violence and mental health outcomes, which can inform state-level policies and intervention programs.^15,17^ Second, socially accepted abuse, such as wife beating, poses a challenge in measuring true state-level estimates of gender-based violence against women.^13^ The National Family Health Survey (2019-2021) revealed that only 8% of IPV victims formally seek help from police, highlighting massive underreporting of crimes against women.^16^ Such reporting biases can mask the true association of state-level GBV with mental health burden, especially as it is likely that there are state-level variations in the extent and nature of underreporting. Third, despite age-standardised disability-adjusted life-years providing a comprehensive measure of disease burden that combines long-term morbidity and premature mortality, this measure is limited by its long-term stability relative to measures of incidence, which would have provided insight into changes in disorder onset and new cases and might be more sensitive to changing rates of GBV.^17^ Fourth, although GBD data enable large-scale, region-specific analyses, they rely on modelled estimates.^26^

The substantial variation in the GBV rates between Indian states calls for geographically targeted action. States with rapidly increasing reported GBV cases may require additional investment in integrated support systems such as One Stop Centres, and capacity building for community mental health.^13^ However, reported GBV rates are influenced by willingness to disclose violence, access to police and support services, and differences in recording practices. Resource allocation should therefore not rely on police-recorded cases alone but should triangulate crime data with population surveys, health service data, and measures of gender inequality. Our findings also shed light on how the public-health implications of GBV are not confined to women. Higher GBV rates were associated with a greater burden of mental health disorders among both women and men, potentially reflecting the impact of wider environments characterised by violence and eco-social stressors.^20^ Prevention strategies should therefore retain a clear focus on protecting women while also addressing harmful cultural and gender norms, perpetrators’ mental-health needs, and the broader social conditions that can contribute to both violence and psychological distress.^28^

## Conclusion

Our state-level findings indicated disorder-specific associations for both sexes, providing comprehensive insights for State governing bodies to address the distribution of mental health disease burden through region-specific policies. Disorder-specific gender disparities can inform intersectoral policy frameworks and advocate for targeted subnational funding to ensure equitable access to health care and social services. Given that the association between gender-based violence and the burden of overall mental health disorders strengthened over time, it is important to address crime against women as a systemic issue with preventive programs such as sex education in schools.

Additionally, the association between increased state-level GBV rates and a rising burden of overall mental health disorders among men over two years underscore the multidimensional impact of gender-based violence that goes beyond women, reflecting a broader societal and structural concern. Our findings therefore provide useful data for researchers and policymakers interested in addressing social determinants of mental health disparities in India.

## Data Availability

All data used in this study are publicly available and can be accessed through the sources cited in the manuscript.

https://www.data.gov.in/

https://ghdx.healthdata.org/

## Contributors

SS conceptualised the study, conducted the literature review, managed data, performed visualisations, formally analysed the data, drafted, and edited the manuscript.

PP conceptualised, designed, supervised the study and reviewed, edited and approved the submission of the manuscript for publication.

GB and JBK reviewed the study design, formal analysis, visualisations, supervised, reviewed, edited, and approved the submission of the manuscript for publication.

All authors had final responsibility for the decision to submit for publication. SS and PP have accessed and verified the data.

## Acknowledgement

We would like to acknowledge that we conducted this research using data from the Global Burden of Diseases, Injuries, and Risk Factors Study (GBD), which is coordinated by the Institute for Health Metrics and Evaluation. The GBD was partially funded by the Bill & Melinda Gates Foundation; the funders had no role in the study design, data analysis, data interpretation, or writing of the report. This project was not developed with consultation or support from the GBD research team.

