## Supplementary material for "Gender-based violence rates and mental health disease burden in India, 2001-2021: A state-level ecological study"

### 1. Supplementary methods

#### A. Gender-based violence rate and mental health disorders estimation for Telangana and Andhra Pradesh

Since the state of Telangana was officially separated from Andhra Pradesh in 2014, the NCRB reported combined crime against women data for Andhra Pradesh and Telangana before 2014. Post 2014, crime cases are reported separately for both States. To maintain consistency and comparability in the dataset over time, we combined the post-2014 crime-against-women data for Andhra Pradesh and Telangana to represent a unified region equivalent to pre-2014 Andhra Pradesh. The combined GBV rate was calculated by:
(a) summing the total number of cases of crime against women for both States;
(b) combining the mid-year populations of Andhra Pradesh and Telangana using weighted outcomes based on population estimates from the National Health Profile 2014.
(c) calculating the GBV rate per 100,000 population using this aggregate mid-year population.
The same method was applied to compute combined age-standardised disability-adjusted life years rates (ASDR) for each of the six mental health outcomes and SDI values. Hence, 27 State clusters were formed by combining Andhra Pradesh & Telangana into a single regional unit. Two UTs were included, resulting in a total of 29 units.

#### B. GBD calculation of age-standarised disability adjusted life years rate (ASDR) for six mental health outcomes with ICD-10 codes.

| Six mental health outcomes | ICD-10 codes |
|---|---|
| 1. Mental disorders | Level 2 (GBD) |
| 2. Schizophrenia | F20 |
| 3. Bipolar disorder | F30.0-F30.9, F31.0-F31.6, F31.8-F31.9, F34.0 |
| 4. Depressive disorders | Major depressive disorder (F32.0-9, F33.0-9), Dysthymia (F34.1) |
| 5. Anxiety disorder | F40-42, F43.0, F43.1, F93.0-93.2, F93.8 |
| 6. Self-harm | X60-X64.9, X66-X84.9, Y87.0 |

GBD calculates DALYs as the sum of years lost due to premature deaths and years lived with disabilities. Formula used by GBD study to calculate DALYs (38):

*DALYs= Results of Years of life lost (YLLs) + Years Lived with disability (YLDs)*

Wherein, YLL = N (number of deaths at age x) x L (standard of life expectancy at age x in years). YLD is measured by multiplying the number of disability cases (i.e., incidence cases) by the average duration of the disease or disability, with a weighted value assigned to the type of disability (disability weight). YLD is measured using: YLD = P (number of prevalent cases) x DW (disability weight)

### 2. Supplementary results

#### A. Descriptive statistics

**Table S1. Descriptive statistics representing age-standardised disability-adjusted life years for depressive disorders among women and men per 100,000 population by State (2001-2021).**

| Sr. No | States | ASDR for depressive disorders among women | | | ASDR for depressive disorders among men | | | Gender disparity ratio |
|---|---|---|---|---|---|---|---|---|
| | | Mean (95% CI) | | | Mean (95% CI) | | | (Female: Male) |
| 1 | Andhra Pradesh & Telangana* | 999·6 | 936·1 | 1063·1 | 793·6 | 749·6 | 837·5 | 1·3 |
| 2 | Arunachal Pradesh | 961·8 | 900·0 | 1023·6 | 744·9 | 703·0 | 786·8 | 1·3 |
| 3 | Assam | 768·2 | 704·8 | 831·7 | 567·0 | 530·3 | 603·8 | 1·4 |
| 4 | Bihar | 731·0 | 688·2 | 773·8 | 545·2 | 515·6 | 574·8 | 1·3 |
| 5 | Chhattisgarh | 626·9 | 594·3 | 659·4 | 430·4 | 402·9 | 457·9 | 1·5 |
| 6 | Delhi** | 678·0 | 636·0 | 720·1 | 528·4 | 498·2 | 558·6 | 1·3 |
| 7 | Goa | 769·9 | 720·6 | 819·2 | 628·1 | 591·9 | 664·3 | 1·2 |
| 8 | Gujarat | 661·0 | 615·3 | 706·7 | 521·1 | 498·7 | 543·5 | 1·3 |
| 9 | Haryana | 1115·9 | 1063·6 | 1168·2 | 781·6 | 750·6 | 812·6 | 1·4 |
| 10 | Himachal Pradesh | 785·0 | 737·5 | 832·6 | 608·5 | 576·8 | 640·2 | 1·3 |
| 11 | Jammu & Kashmir** | 735·6 | 691·2 | 780·0 | 585·6 | 553·1 | 618·2 | 1·3 |
| 12 | Jharkhand | 713·4 | 688·7 | 738·1 | 643·9 | 630·4 | 657·4 | 1·1 |
| 13 | Karnataka | 747·2 | 706·0 | 788·4 | 581·5 | 554·7 | 608·3 | 1·3 |
| 14 | Kerala | 756·7 | 722·2 | 791·2 | 614·9 | 583·7 | 646·2 | 1·2 |
| 15 | Madhya Pradesh | 708·9 | 662·9 | 754·9 | 471·0 | 439·9 | 502·1 | 1·5 |
| 16 | Maharashtra | 982·8 | 933·0 | 1032·5 | 728·9 | 700·6 | 757·2 | 1·3 |
| 17 | Manipur | 954·9 | 918·3 | 991·5 | 614·2 | 589·5 | 638·9 | 1·6 |
| 18 | Meghalaya | 913·5 | 857·9 | 969·2 | 733·9 | 692·8 | 775·0 | 1·2 |
| 19 | Mizoram | 705·8 | 663·8 | 747·8 | 560·3 | 528·8 | 591·8 | 1·3 |
| 20 | Nagaland | 790·0 | 746·2 | 833·8 | 623·5 | 591·0 | 655·9 | 1·3 |
| 21 | Odisha | 978·2 | 916·4 | 1040·0 | 758·5 | 716·1 | 801·0 | 1·3 |
| 22 | Punjab | 685·1 | 660·9 | 709·3 | 448·4 | 426·6 | 470·3 | 1·5 |
| 23 | Rajasthan | 679·1 | 655·6 | 702·6 | 546·5 | 533·1 | 559·9 | 1·2 |
| 24 | Sikkim | 794·8 | 751·0 | 838·6 | 605·6 | 573·0 | 638·3 | 1·3 |
| 25 | Tamil Nadu | 1189·8 | 1146·0 | 1233·6 | 790·6 | 764·9 | 816·2 | 1·5 |
| 26 | Tripura | 739·6 | 694·0 | 785·2 | 580·2 | 546·9 | 613·4 | 1·3 |
| 27 | Uttar Pradesh | 671·9 | 635·2 | 708·6 | 497·2 | 479·4 | 515·1 | 1·4 |
| 28 | Uttarakhand | 726·0 | 681·9 | 770·0 | 571·3 | 538·8 | 603·7 | 1·3 |
| 29 | West Bengal | 833·4 | 809·8 | 857·0 | 509·1 | 496·4 | 521·8 | 1·6 |

Note: The descriptive statistics represent an average ASDR for depressive disorders over 21 years (2001–2021); 95%CI= 95% confidence interval; GBV rates= gender-based incidence rates, ASDR= age standardised disability-adjusted life years

**Union territories considered for analysis.

*Combined two separate States for this analysis.

**Table S2. Descriptive statistics representing average age-standardised disability-adjusted life years for anxiety disorders among women and men per 100,000 population by State (2001-2021).**

| Sr. No | States | ASDR for anxiety disorders among women | | | ASDR for anxiety disorders among men | | | Gender disparity ratio |
|---|---|---|---|---|---|---|---|---|
| | | Mean (95% CI) | | | Mean (95% CI) | | | (Female: Male) |
| 1 | Andhra Pradesh & Telangana* | 425·8 | 414·8 | 436·7 | 293·0 | 282·0 | 304·0 | 1·5 |
| 2 | Arunachal Pradesh | 426·0 | 418·3 | 433·8 | 293·0 | 283·8 | 302·2 | 1·5 |
| 3 | Assam | 426·1 | 415·0 | 437·1 | 293·2 | 282·3 | 304·0 | 1·5 |
| 4 | Bihar | 425·0 | 416·2 | 433·8 | 292·8 | 283·0 | 302·5 | 1·5 |
| 5 | Chhattisgarh | 299·6 | 290·2 | 308·9 | 193·3 | 187·8 | 198·7 | 1·6 |
| 6 | Delhi** | 428·4 | 415·1 | 441·7 | 295·2 | 283·1 | 307·2 | 1·5 |
| 7 | Goa | 297·4 | 287·7 | 307·1 | 207·8 | 201·7 | 213·8 | 1·4 |
| 8 | Gujarat | 390·1 | 380·7 | 399·4 | 235·1 | 228·1 | 242·1 | 1·7 |
| 9 | Haryana | 427·7 | 414·9 | 440·5 | 294·0 | 282·3 | 305·8 | 1·5 |
| 10 | Himachal Pradesh | 429·0 | 418·0 | 440·0 | 293·9 | 283·2 | 304·6 | 1·5 |
| 11 | Jammu & Kashmir** | 429·0 | 418·1 | 439·8 | 294·3 | 283·7 | 305·0 | 1·5 |
| 12 | Jharkhand | 474·5 | 462·4 | 486·6 | 360·5 | 352·1 | 369·0 | 1·3 |
| 13 | Karnataka | 429·3 | 416·2 | 442·3 | 294·1 | 282·2 | 305·9 | 1·5 |
| 14 | Kerala | 568·1 | 536·2 | 600·0 | 352·9 | 338·7 | 367·2 | 1·6 |
| 15 | Madhya Pradesh | 283·5 | 277·1 | 289·9 | 203·4 | 198·7 | 208·1 | 1·4 |
| 16 | Maharashtra | 431·7 | 415·2 | 448·1 | 296·3 | 282·5 | 310·1 | 1·5 |
| 17 | Manipur | 590·8 | 567·5 | 614·2 | 381·9 | 368·2 | 395·6 | 1·5 |
| 18 | Meghalaya | 426·1 | 415·5 | 436·7 | 292·9 | 282·3 | 303·4 | 1·5 |
| 19 | Mizoram | 430·3 | 418·5 | 442·1 | 294·7 | 283·6 | 305·8 | 1·5 |
| 20 | Nagaland | 430·1 | 419·9 | 440·4 | 293·6 | 283·4 | 303·9 | 1·5 |
| 21 | Odisha | 425·4 | 415·1 | 435·6 | 292·5 | 282·0 | 303·0 | 1·5 |
| 22 | Punjab | 373·8 | 363·1 | 384·6 | 262·4 | 254·1 | 270·8 | 1·4 |
| 23 | Rajasthan | 455·6 | 444·1 | 467·1 | 340·9 | 333·0 | 348·8 | 1·3 |
| 24 | Sikkim | 425·3 | 417·1 | 433·6 | 292·9 | 283·6 | 302·2 | 1·5 |
| 25 | Tamil Nadu | 422·6 | 410·2 | 435·0 | 264·6 | 255·2 | 274·0 | 1·6 |
| 26 | Tripura | 426·4 | 417·1 | 435·8 | 293·7 | 283·8 | 303·6 | 1·5 |
| 27 | Uttar Pradesh | 425·8 | 415·0 | 436·7 | 256·4 | 250·2 | 262·5 | 1·7 |
| 28 | Uttarakhand | 427·7 | 416·5 | 439·0 | 293·0 | 282·1 | 304·0 | 1·5 |
| 29 | West Bengal | 483·7 | 469·0 | 498·4 | 280·3 | 272·4 | 288·3 | 1·7 |

Note: The descriptive statistics represent an average ASDR for anxiety disorders over 21 years (2001–2021); 95%CI= 95% confidence interval; GBV rates= gender-based incidence rates; ASDR = age-standardised disability-adjusted life years

**Union territories considered for analysis.

*Combined two separate States for this analysis.

**Table S3. Descriptive statistics representing the average age-standardised disability-adjusted life years for schizophrenia among women and men per 100,000 population by State (2001-2021).**

| States | ASDR for schizophrenia among women | | | ASDR for schizophrenia among men | | | Gender disparity ratio |
|---|---|---|---|---|---|---|---|
| | Mean (95% CI) | | | Mean (95% CI) | | | (Female: Male) |
| Andhra Pradesh & Telangana* | 158·2 | 156·4 | 160·1 | 210·6 | 208·8 | 212·5 | 0·75 |
| Arunachal Pradesh | 159·8 | 158·1 | 161·6 | 212·3 | 210·6 | 213·9 | 0·75 |
| Assam | 157·0 | 155·2 | 158·7 | 209·1 | 207·6 | 210·6 | 0·75 |
| Bihar | 157·0 | 155·2 | 158·7 | 209·7 | 208·1 | 211·2 | 0·75 |
| Chhattisgarh | 157·8 | 156·0 | 159·7 | 209·7 | 208·2 | 211·2 | 0·75 |
| Delhi** | 160·4 | 158·5 | 162·3 | 213·7 | 212·0 | 215·3 | 0·75 |
| Goa | 163·0 | 161·2 | 164·8 | 216·5 | 215·0 | 217·9 | 0·75 |
| Gujarat | 158·0 | 156·3 | 159·7 | 211·6 | 210·1 | 213·1 | 0·75 |
| Haryana | 159·6 | 157·8 | 161·4 | 211·3 | 209·6 | 213·0 | 0·76 |
| Himachal Pradesh | 161·4 | 159·6 | 163·2 | 212·9 | 211·4 | 214·5 | 0·76 |
| Jammu & Kashmir** | 160·5 | 158·8 | 162·3 | 212·8 | 211·4 | 214·2 | 0·75 |
| Jharkhand | 157·1 | 155·5 | 158·7 | 210·3 | 208·7 | 212·0 | 0·75 |
| Karnataka | 159·3 | 157·6 | 161·0 | 210·8 | 209·1 | 212·5 | 0·76 |
| Kerala | 162·9 | 161·1 | 164·7 | 214·0 | 212·3 | 215·7 | 0·76 |
| Madhya Pradesh | 158·1 | 156·4 | 159·8 | 209·5 | 208·1 | 211·0 | 0·75 |
| Maharashtra | 159·9 | 158·0 | 161·7 | 211·8 | 210·2 | 213·5 | 0·75 |
| Manipur | 159·6 | 158·0 | 161·2 | 211·3 | 209·9 | 212·6 | 0·75 |
| Meghalaya | 159·0 | 157·2 | 160·8 | 211·1 | 209·6 | 212·6 | 0·75 |
| Mizoram | 160·7 | 158·8 | 162·5 | 212·9 | 211·3 | 214·4 | 0·75 |
| Nagaland | 160·7 | 159·0 | 162·5 | 212·3 | 210·7 | 213·8 | 0·76 |
| Odisha | 157·1 | 155·3 | 158·9 | 208·7 | 207·1 | 210·4 | 0·75 |
| Punjab | 161·0 | 159·3 | 162·7 | 212·4 | 210·7 | 214·1 | 0·76 |
| Rajasthan | 159·6 | 157·9 | 161·3 | 211·5 | 210·1 | 213·0 | 0·75 |
| Sikkim | 160·6 | 158·6 | 162·5 | 213·8 | 211·9 | 215·7 | 0·75 |
| Tamil Nadu | 146·7 | 146·4 | 146·9 | 198·4 | 198·1 | 198·8 | 0·74 |
| Tripura | 159·3 | 157·5 | 161·0 | 211·9 | 210·5 | 213·3 | 0·75 |
| Uttar Pradesh | 157·4 | 155·8 | 159·1 | 208·9 | 207·4 | 210·3 | 0·75 |
| Uttarakhand | 158·9 | 157·0 | 160·9 | 209·5 | 207·8 | 211·2 | 0·76 |
| West Bengal | 158·4 | 156·7 | 160·2 | 212·1 | 210·5 | 213·7 | 0·75 |

Note: The descriptive statistics represent an average ASDR for schizophrenia over 21 years (2001–2021); 95%CI= 95% confidence interval; GBV rates= gender-based incidence rates, ASDR= age standardised disability-adjusted life years

**Union territories considered for analysis.

*Combined two separate States for this analysis.

**Table S4. Descriptive statistics representing the average age-standardised disability-adjusted life years for bipolar disorder among women and men per 100,000 population by State (2001-2021).**

| **States** | **ASDR for bipolar disorder among women** | | | **ASDR for bipolar disorder among men** | | | **Gender disparity ratio** |
|---|---|---|---|---|---|---|---|
| | **Mean (95% CI)** | | | **Mean (95% CI)** | | | **(Female: Male)** |
| Andhra Pradesh & Telangana* | 64·1 | 64·0 | 64·1 | 75·5 | 75·4 | 75·6 | 0·85 |
| Arunachal Pradesh | 64·3 | 64·3 | 64·4 | 75·7 | 75·7 | 75·8 | 0·85 |
| Assam | 63·9 | 63·8 | 64·0 | 75·3 | 75·2 | 75·4 | 0·85 |
| Bihar | 64·0 | 63·9 | 64·1 | 75·5 | 75·4 | 75·6 | 0·85 |
| Chhattisgarh | 64·2 | 64·1 | 64·2 | 75·4 | 75·3 | 75·4 | 0·85 |
| Delhi** | 64·2 | 64·1 | 64·3 | 75·9 | 75·8 | 75·9 | 0·85 |
| Goa | 65·0 | 64·9 | 65·0 | 76·3 | 76·2 | 76·3 | 0·85 |
| Gujarat | 63·6 | 63·6 | 63·7 | 75·7 | 75·6 | 75·7 | 0·84 |
| Haryana | 64·1 | 64·0 | 64·1 | 75·5 | 75·5 | 75·6 | 0·85 |
| Himachal Pradesh | 64·9 | 64·8 | 64·9 | 76·0 | 75·9 | 76·0 | 0·85 |
| Jammu & Kashmir** | 64·7 | 64·6 | 64·7 | 75·9 | 75·9 | 76·0 | 0·85 |
| Jharkhand | 63·8 | 63·8 | 63·9 | 75·6 | 75·5 | 75·7 | 0·84 |
| Karnataka | 64·3 | 64·3 | 64·4 | 75·6 | 75·6 | 75·7 | 0·85 |
| Kerala | 64·9 | 64·8 | 64·9 | 76·0 | 76·0 | 76·1 | 0·85 |
| Madhya Pradesh | 64·1 | 64·1 | 64·2 | 75·5 | 75·5 | 75·6 | 0·85 |
| Maharashtra | 64·4 | 64·3 | 64·5 | 75·7 | 75·6 | 75·7 | 0·85 |
| Manipur | 64·6 | 64·5 | 64·6 | 75·8 | 75·8 | 75·9 | 0·85 |
| Meghalaya | 64·1 | 64·1 | 64·2 | 75·6 | 75·5 | 75·7 | 0·85 |
| Mizoram | 64·7 | 64·7 | 64·8 | 75·9 | 75·8 | 75·9 | 0·85 |
| Nagaland | 64·8 | 64·7 | 64·8 | 75·9 | 75·8 | 76·0 | 0·85 |
| Odisha | 63·9 | 63·8 | 64·0 | 75·3 | 75·2 | 75·4 | 0·85 |
| Punjab | 64·6 | 64·6 | 64·7 | 75·8 | 75·7 | 75·9 | 0·85 |
| Rajasthan | 64·4 | 64·4 | 64·5 | 75·9 | 75·8 | 75·9 | 0·85 |
| Sikkim | 64·4 | 64·3 | 64·5 | 75·9 | 75·8 | 76·0 | 0·85 |
| Tamil Nadu | 53·0 | 52·8 | 53·1 | 62·9 | 62·7 | 63·0 | 0·84 |
| Tripura | 64·4 | 64·3 | 64·4 | 75·7 | 75·7 | 75·8 | 0·85 |
| Uttar Pradesh | 64·0 | 64·0 | 64·1 | 75·3 | 75·2 | 75·4 | 0·85 |
| Uttarakhand | 64·2 | 64·1 | 64·2 | 75·4 | 75·3 | 75·5 | 0·85 |
| West Bengal | 63·9 | 63·9 | 64·0 | 75·7 | 75·6 | 75·8 | 0·84 |

Note: The descriptive statistics represent an average ASDR for bipolar disorder over 21 years (2001–2021); 95%CI= 95% confidence interval; GBV rates= gender-based incidence rates; ASDR = age-standardised disability-adjusted life years
**Union territories considered for analysis.
*Combined two separate States for this analysis.

**Table S5. Descriptive statistics representing average age-standardised disability-adjusted life year rate for self-harm among women and men per 100,000 population by State (2001-2021).**

| States | ASDR for self-harm among women | | | ASDR for self-harm among men | | | Gender disparity ratio |
|---|---|---|---|---|---|---|---|
| | Mean (95% CI) | | | Mean (95% CI) | | | (Female: Male) |
| Andhra Pradesh & Telangana* | 1034·7 | 889·7 | 1179·7 | 1230·5 | 1145·6 | 1315·3 | 0·84 |
| Arunachal Pradesh | 708·2 | 638·9 | 777·4 | 822·6 | 773·9 | 871·2 | 0·87 |
| Assam | 750·9 | 639·9 | 861·9 | 790·2 | 732·1 | 848·4 | 0·95 |
| Bihar | 307·5 | 289·5 | 325·5 | 313·5 | 306·7 | 320·2 | 1·00 |
| Chhattisgarh | 707·9 | 654·3 | 761·5 | 1457·4 | 1411·1 | 1503·7 | 0·50 |
| Delhi** | 299·6 | 268·3 | 330·9 | 378·8 | 356·4 | 401·2 | 0·80 |
| Goa | 619·4 | 550·9 | 687·9 | 513·4 | 477·1 | 549·7 | 1·21 |
| Gujarat | 801·3 | 761·1 | 841·5 | 806·0 | 772·5 | 839·5 | 1·00 |
| Haryana | 543·9 | 500·5 | 587·3 | 813·8 | 757·4 | 870·1 | 0·67 |
| Himachal Pradesh | 547·6 | 493·5 | 601·8 | 802·4 | 765·8 | 838·9 | 0·70 |
| Jammu & Kashmir** | 378·1 | 354·1 | 402·1 | 276·0 | 261·5 | 290·5 | 1·39 |
| Jharkhand | 445·4 | 404·6 | 486·3 | 424·2 | 368·3 | 480·0 | 1·06 |
| Karnataka | 1107·7 | 1034·9 | 1180·4 | 1536·8 | 1452·6 | 1621·0 | 0·72 |
| Kerala | 604·9 | 555·3 | 654·4 | 1311·2 | 1219·0 | 1403·3 | 0·46 |
| Madhya Pradesh | 809·8 | 745·5 | 874·2 | 988·4 | 965·6 | 1011·3 | 0·84 |
| Maharashtra | 588·5 | 542·3 | 634·7 | 847·9 | 809·8 | 886·0 | 0·69 |
| Manipur | 426·3 | 393·3 | 459·3 | 703·4 | 683·8 | 723·1 | 0·61 |
| Meghalaya | 194·7 | 178·1 | 211·2 | 548·0 | 524·2 | 571·7 | 0·36 |
| Mizoram | 170·3 | 149·2 | 191·3 | 442·3 | 419·2 | 465·4 | 0·39 |
| Nagaland | 155·1 | 134·7 | 175·4 | 249·2 | 244·7 | 253·6 | 0·64 |
| Odisha | 782·7 | 700·1 | 865·4 | 795·7 | 754·1 | 837·3 | 0·99 |
| Punjab | 365·3 | 347·8 | 382·8 | 476·0 | 452·6 | 499·3 | 0·78 |
| Rajasthan | 392·1 | 376·4 | 407·8 | 614·9 | 597·3 | 632·4 | 0·64 |
| Sikkim | 534·6 | 456·8 | 612·4 | 711·5 | 652·7 | 770·4 | 0·75 |
| Tamil Nadu | 1603·0 | 1427·6 | 1778·4 | 1653·7 | 1553·5 | 1753·9 | 0·97 |
| Tripura | 970·9 | 863·3 | 1078·5 | 1607·4 | 1525·9 | 1688·9 | 0·61 |
| Uttar Pradesh | 705·3 | 661·1 | 749·5 | 684·8 | 660·2 | 709·4 | 1·03 |
| Uttarakhand | 800·9 | 717·6 | 884·2 | 877·7 | 830·8 | 924·7 | 0·92 |
| West Bengal | 913·0 | 836·9 | 989·2 | 920·6 | 870·9 | 970·3 | 0·99 |

Note: The descriptive statistics represent an average ASDR for self-harm over 21 years (2001–2021); 95%CI= 95% confidence interval; GBV rates= gender-based incidence rates; ASDR = age-standardised disability-adjusted life years
**Union territories considered for analysis.
*Combined two separate States for this analysis

**B. Intraclass correlation coefficients of mental health disorders, by gender, 2001-2021**

**Table S6. Empty Model Intraclass Correlation Coefficients (ICC) of ASDR for six mental health disorders among women, men and gender disparity ratio.**

| **Outcome** | **Null model** | | |
|---|---|---|---|
| | **women** | **men** | **Gender disparity ratio** |
| | Intraclass Correlation Coefficients (ICC) | | |
| Mental health disorders | 0·73 | 0·79 | 0·85 |
| Depressive disorders | 0·66 | 0·69 | 0·92 |
| Anxiety disorders | 0·82 | 0·77 | 0·75 |
| Self-harm | 0·80 | 0·93 | 0·83 |
| Bipolar disorder | 1·00 | 1·00 | 0·75 |
| Schizophrenia | 0·33 | 0·41 | 0·31 |

Intraclass correlation coefficient (ICC) indicates that a substantial proportion of the variance in overall mental health outcomes–including anxiety disorders (women ICC=0.82, men ICC=0.77), depressive disorders (women ICC=0.66, men ICC=0.69), bipolar disorder (women ICC=1, men ICC=1) and self-harm (women ICC=0.80, men ICC=0.93) was attributable to between-state differences among women and men. A potential explanation could be that regional factors such as healthcare infrastructure, socioeconomic conditions, and resource availability strongly influence the early detection of these disorders, leading to greater clustering within States. On the contrary, schizophrenia showed the lowest between-state variance (women ICC = 0.33; men ICC = 0.41), indicating a less regionally clustered pattern (Table S6).

### C. Sensitivity Analyses

**Table S7. Sensitivity analyses for the fixed effects model with a time lag of one year (t-1) and five years (t-5) from 2001 to 2021**

| Outcome | Fixed effects model (lag t-1) | | | Fixed effects model (lag t-5) | | |
|---|---|---|---|---|---|---|
| | women | men | **Gender disparity rate ratio** | women | men | **Gender disparity rate ratio** |
| GBV rate as exposure, Estimate; 95% CI=LL, UL | | | | | | |
| Mental health disorders | 6·19 (-13·43, 25·81) | 5·51 (-5·94, 16·95) | -0·00 (-0·00,0·00) | 19·19* (4·02, 34·36) | 14·49** (3·92, 25·06) | 0·00 (-0·00, 0·01) |
| Depressive Disorders | 3·72 (-12·49, 19·94) | 3·62 (-7·14, 14·39) | -0·00 (-0·01, 0·00) | 14·45* (0·78, 28·13) | 11·23* (1·09, 21·38) | -0·00 (-0·01, 0·00) |
| Anxiety Disorder | 0·71 (-2·77, 4·20) | 0·22 (-1·04, 1·49) | 0·00 (-0·01, 0·01) | 2·74 (-0·88, 6·35) | 1·83 (-0·65, 4·30) | 0·00 (-0·01, 0·01) |
| Self-harm | -7·46 (-39·22, 24·30) | -8·26 (-25·55, 9·03) | -0·00 (-0·02, 0·01) | -11·37 (-49·75, 27·01) | -9·48 (-34·72, 15·77) | -0·02* (-0·03, 0·00) |
| Bipolar Disorder | 0·02 (-0·01, 0·05) | 0·04* (0·01, 0·07) | -0·00 (-0·00, 0·00) | 0·03 (-0·01, 0·06) | -0·00 (-0·04, 0·04) | 0·00 (-0·00, 0·00) |
| Schizophrenia | 0·44* (0·05, 0·82) | 0·46* (0·05, 0·87) | 0·00* (0·00, 0·00) | 0·12 (-0·41, 0·64) | 0·09 (-0·47, 0·65) | 0·00 (-0·00, 0·00) |

Note: estimate=coefficient, *p<0.05; **p<0.01, ***p<0.001; 95% CI= 95% confidence interval; LL=lower limit, UL=upper limit, GBV rates= gender-based violence incidence rates, SDI=socio-demographic index values. Note: The GBV rate is rescaled to 10,000 population.

**Table S8. COVID-19 pandemic sensitivity analysis from 2001 to 2019**

| Outcome | Mixed effects unadjusted model | | | Mixed effects adjusted model for SDI | | | Fixed effects model (lag t-2) | | |
|---|---|---|---|---|---|---|---|---|---|
| | women | men | Gender disparity rate ratio | women | men | Gender disparity ratio | women | men | Gender disparity rate ratio |
| GBV rate as exposure, Estimate; 95% CI=LL, UL | | | | | | | | | |
| Mental health disorders | 3·80 (-1·82, 9·42) | 4·96* (1·04, 8·88) | -0·00 (-0·02, 0·00) | 3·72 (-1·91, 9·35) | 5·02* (1·10, 8·95) | -0·00 (-0·00, 0·00) | 5·29 (-8·78, 19·36) | 5·68 (-3·35, 14·72) | -0·00 (-0·00, 0·00) |
| Depressive disorders | 2·99 (-2·63, 8·62) | 3·53 (-0·75, 7·81) | -0·004** (-0·007, -0·001) | 3·18 (-2·45, 8·81) | 3·72 (-0·56, 7·99) | -0·005** (-0·008, -0·002) | 3·60 (-10·48, 17·67) | 3·98 (-5·91, 13·87) | -0·01 (-0·01, 0·00) |
| Anxiety disorder | -0·60 (-1·84, 0·65) | -0·29 (-1·09, 0·50) | 0·00 (-0·00, 0·00) | -0·72 (1·95, 0·51) | -0·31 (-1·10, 0·49) | 0·00 (-0·00, 0·00) | -0·27 (-1·83, 1·30) | -0·09 (-1·79, 1·62) | 0·00 (-0·01, 0·01) |
| Self-harm | -5·87 (-15·55, 3·81) | -5·65 (-13·67, 2·37) | -0·00 (-0·01, 0·01) | -6·49 (-16·13, 3·15) | -6·51 (-14·42, 1·41) | -0·00 (-0·01, 0·01) | -7·29 (-43·21, 28·64) | -12·65 (-32·85, 7·54) | -0·01 (-0·02, 0·01) |
| Bipolar disorder | 0·03*** (0·02,0·04) | 0·04*** (0·02, 0·05) | -0·00 (-0·00, 0·00) | 0·03*** (0·02, 0·04) | 0·04*** (0·02, 0·05) | -0·00 (-0·00, 0·00) | 0·03 (-0·01, 0·07) | 0·04 (-0·00, 0·07) | 0·00 (-0·00, 0·00) |
| Schizophrenia | 0·49*** (0·33,0·65) | 0·53*** (0·37, 0·69) | 0·0004** (0·0002, 0·0007) | 0·52*** (0·36, 0·67) | 0·56*** (0·41, 0·71) | 0·0005** (0·0002, 0·0008) | 0·59** (0·21, 0·97) | 0·57* (0·14, 1·00) | 0·0007*** (0·0004, 0·0012) |

*Note: estimate=coefficient, *p<0.05; **p<0.01, ***p<0.001; 95% CI= 95% confidence interval; LL=lower limit, UL=upper limit, GBV rates= gender-based violence incidence rates, SDI=socio-demographic index values.* Note: The GBV rate is rescaled to 10,000 population

### 3. Supplementary figures

**Figure S1.1 Scatterplot panels showing the disorder-specific relationships between mean gender-based violence rates and mean age-standardised disability rates for women across 30 geographical units (Indian states/Union Territories) over time (2001-2021).**

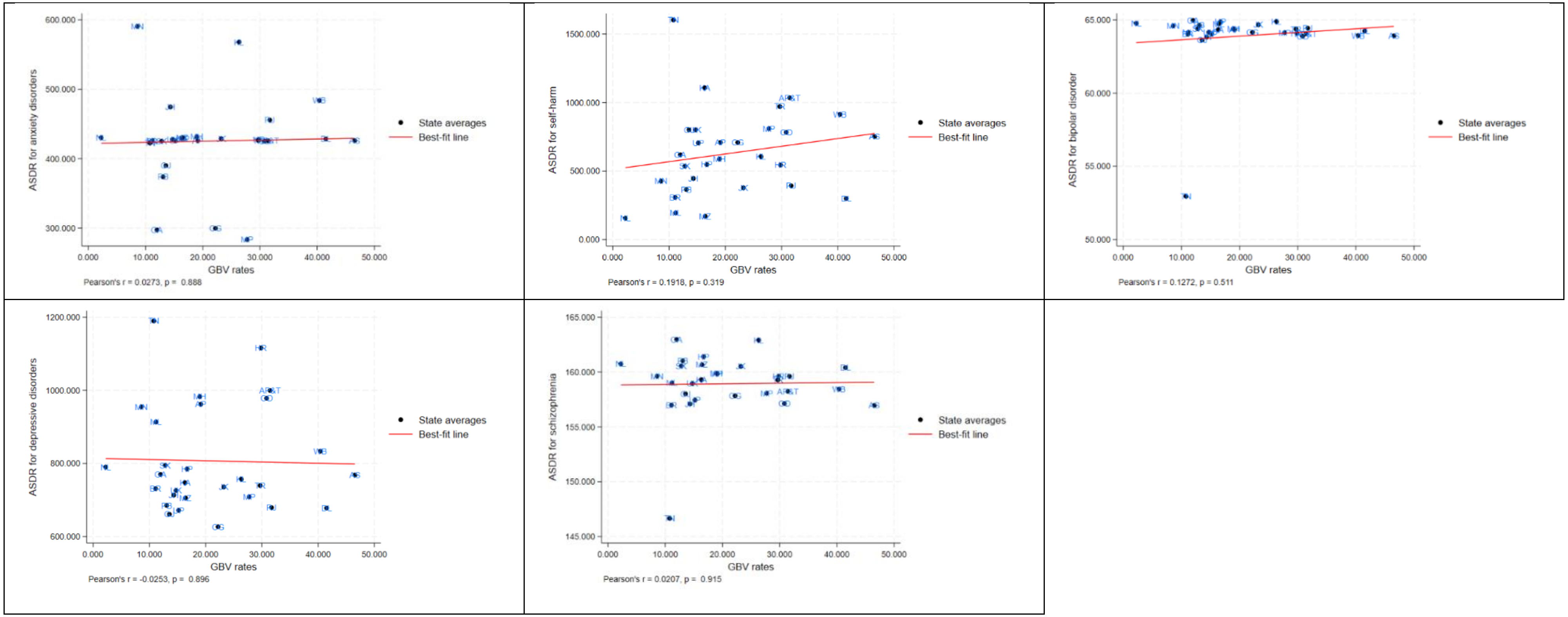

Note. The red line represents the line of best fit, illustrating the trend and strength of correlation between gender-based violence rates and mental health outcomes among women. The data correlated is a cumulative mean of 21 years. Each dot represents the following 30 geographical units (Indian states/Union territories)- Andhra Pradesh & Telangana= AP&T, Arunachal Pradesh=AP, Assam=AS, Bihar=BR, Chhattisgarh=CG, Delhi=DL, Goa=GA, Gujarat=GJ, Haryana=HR, Himachal Pradesh=HP, Jammu & Kashmir=JK, Jharkhand=JH, Karnataka=KA, Kerala=KL, Madhya Pradesh=MP, Maharashtra=MH, Manipur=MN, Meghalaya=ML, Mizoram=MZ, Nagaland=NL, Odisha=OD, Punjab=PB, Rajasthan=RJ, Sikkim=SK, Tamil Nadu=TN, Tripura=TR, Uttar Pradesh=UP, Uttarakhand=UK; West Bengal=WB.

**Figure S1.2 Scatterplot panels showing the disorder-specific relationships between mean gender-based violence rates and mean age-standardised disability rates for men across 30 geographical units (Indian states/Union Territories) over time (2001-2021).**

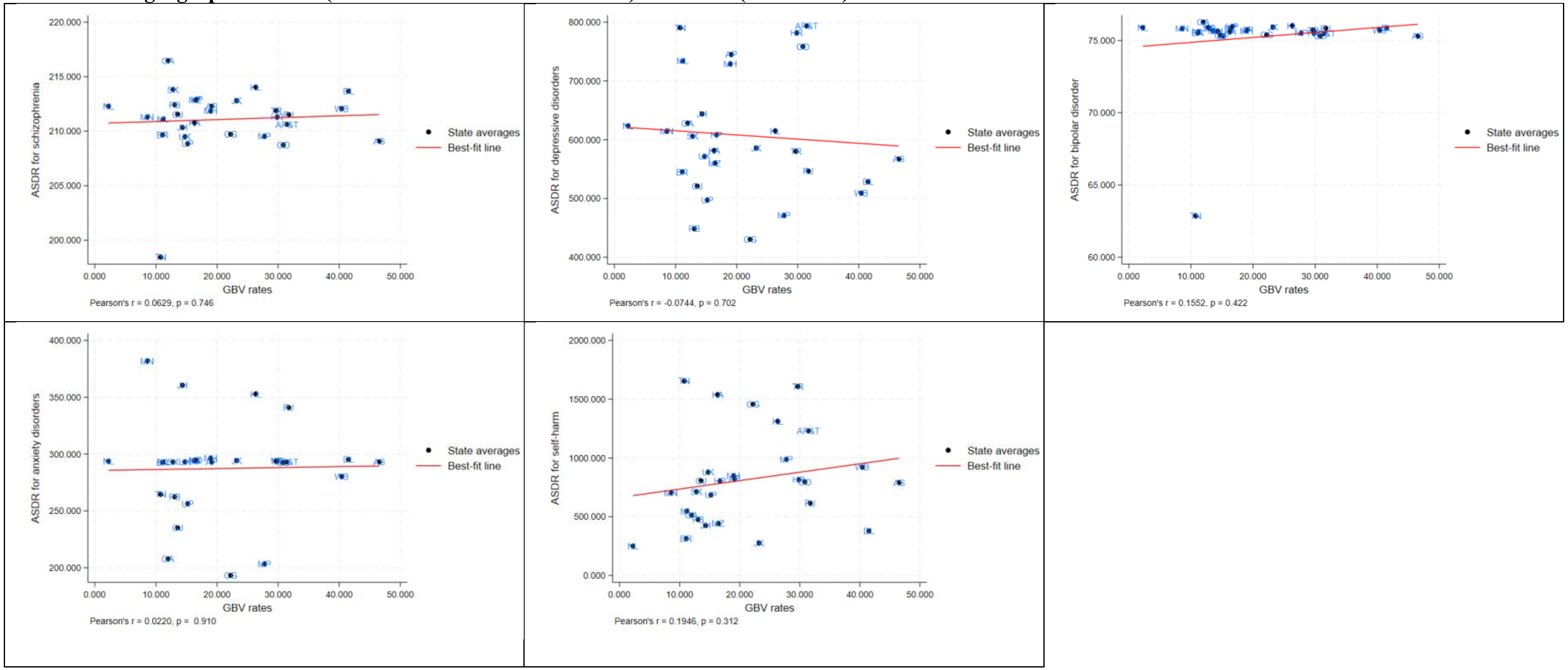

Note. Each point represents a State. The red line represents the line of best fit, illustrating the trend and strength of correlation between GBV rates and ASDR for mental health outcomes among men. The data correlated is a cumulative mean of 21 years. Andhra Pradesh & Telangana= AP&T, Arunachal Pradesh=AP, Assam=AS, Bihar=BR, Chhattisgarh=CG, Delhi=DL, Goa=GA, Gujarat=GJ, Haryana=HR, Himachal Pradesh=HP, Jammu & Kashmir=JK, Jharkhand=JH, Karnataka=KA, Kerala=KL, Madhya Pradesh=MP, Maharashtra=MH, Manipur=MN, Meghalaya=ML, Mizoram=MZ, Nagaland=NL, Odisha=OD, Punjab=PB, Rajasthan=RJ, Sikkim=SK, Tamil Nadu=TN, Tripura=TR, Uttar Pradesh=UP, Uttarakhand=UK; West Bengal=WB.

**Figure S1.3 Scatterplot showing relationships between mean gender-based violence rates and gender disparity ratios of six mental health outcomes across 30 geographical units (Indian states/Union Territories) over time (2001-2021).**

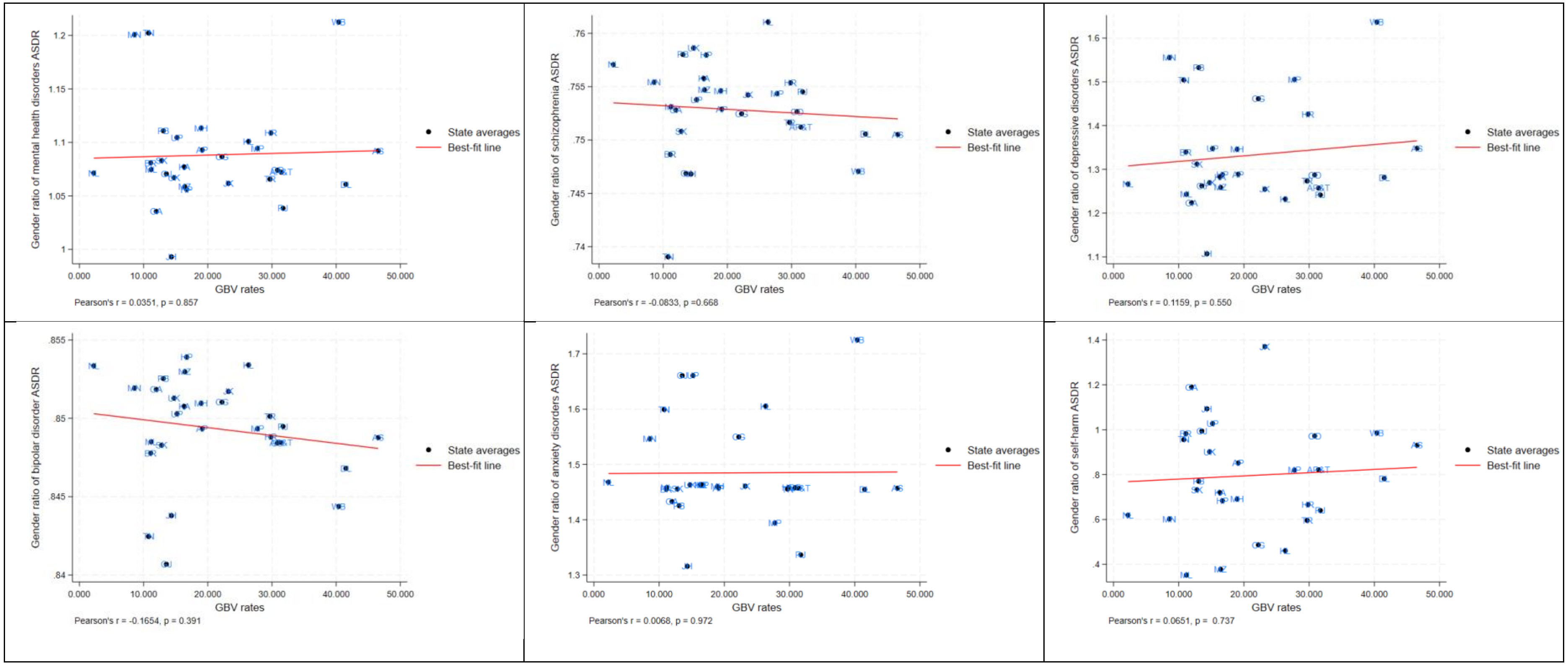

Note. Each point represents a State. The red line represents the line of best fit, illustrating the trend and strength of correlation between GBV rates and gender disparity ratios of each disorder. The data correlated is a cumulative mean of 21 years. Andhra Pradesh & Telangana= AP&T, Arunachal Pradesh=AP, Assam=AS, Bihar=BR, Chhattisgarh=CG, Delhi=DL, Goa=GA, Gujarat=GJ, Haryana=HR, Himachal Pradesh=HP, Jammu & Kashmir=JK, Jharkhand=JH, Karnataka=KA, Kerala=KL, Madhya Pradesh=MP, Maharashtra=MH, Manipur=MN, Meghalaya=ML, Mizoram=MZ, Nagaland=NL, Odisha=OD, Punjab=PB, Rajasthan=RJ, Sikkim=SK, Tamil Nadu=TN, Tripura=TR, Uttar Pradesh=UP, Uttarakhand=UK; West Bengal=WB.

**Figure S2.1 Distribution of gender-based violence rates and gender disparity in depressive disorders burden across 30 geographical units (Indian states/ Union Territories) at two time points, 2001 and 2021.**

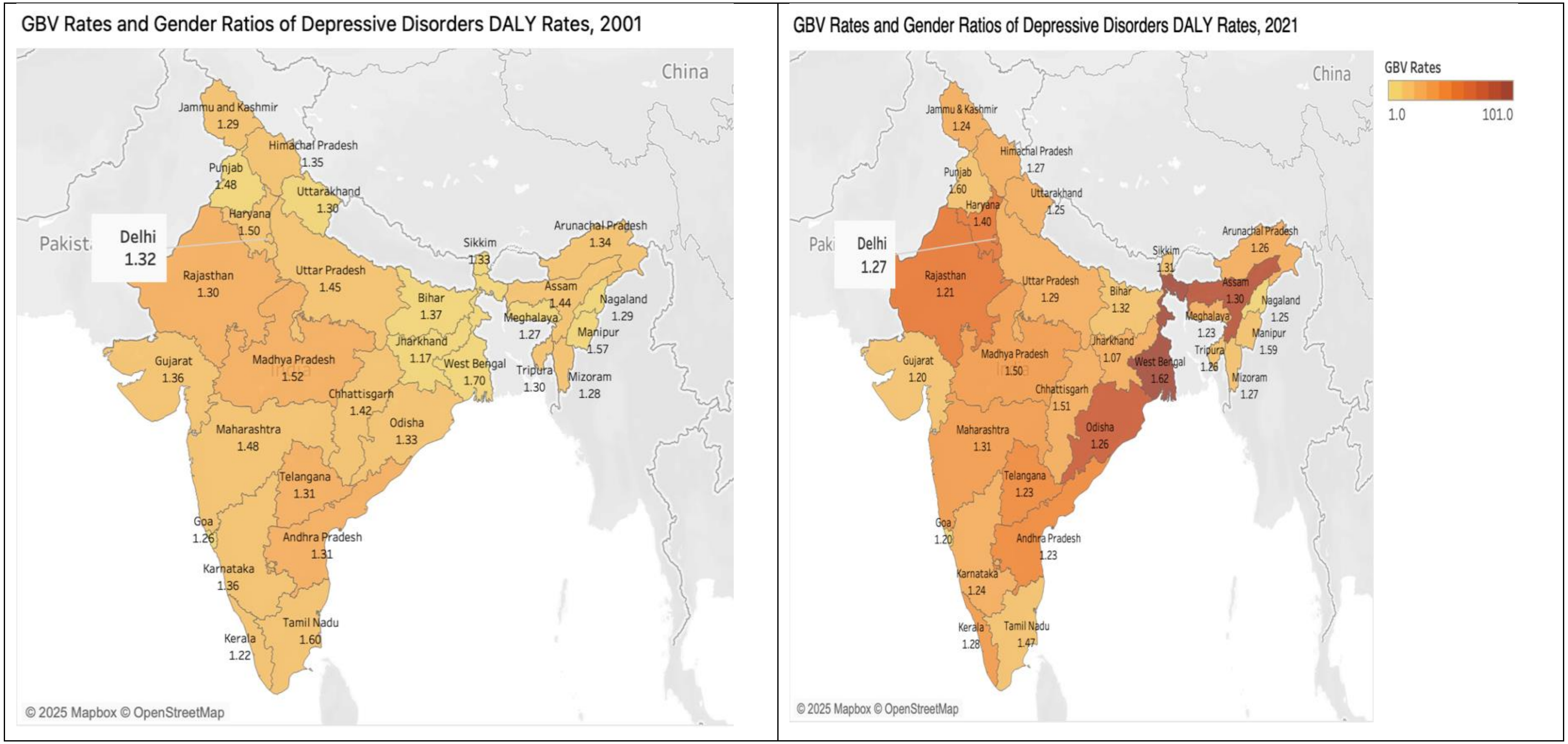

Note: The Indian map displays gender-based violence rates by colour gradient, with darker shades denoting higher rates and lighter shades denoting lower rates. Each geographical unit is labelled with the gender disparity ratio of depressive disorders burden at baseline (2001) and the most recent year (2021). A gender disparity ratio greater than 1 denotes a higher depressive disorders burden among women than men per 100,000 population.

**Figure S2.2 Distribution of gender-based violence rates and gender disparity in anxiety disorders burden across 30 geographical units (Indian states/ Union Territories) at two time points, 2001 and 2021.**

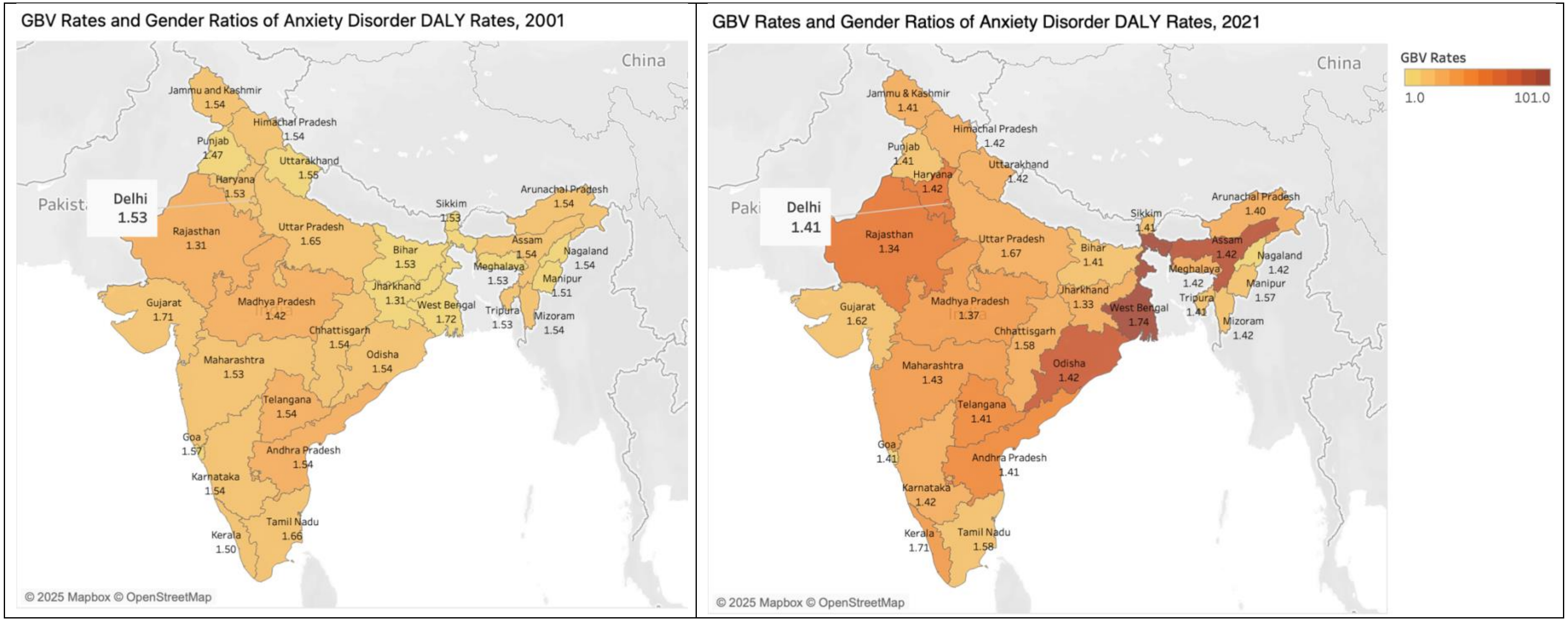

Note: The Indian map displays gender-based violence rates by colour gradient, with darker shades denoting higher rates and lighter shades denoting lower rates. Each geographical unit is labelled with the gender disparity ratio of anxiety disorders burden at baseline (2001) and the most recent year (2021). A gender disparity ratio greater than 1 denotes a higher anxiety disorders burden among women than men per 100,000 population.

**Figure S2.3 Distribution of gender-based violence rates and gender disparity in bipolar disorder burden across 30 geographical units (Indian states/ Union Territories) at two time points, 2001 and 2021.**

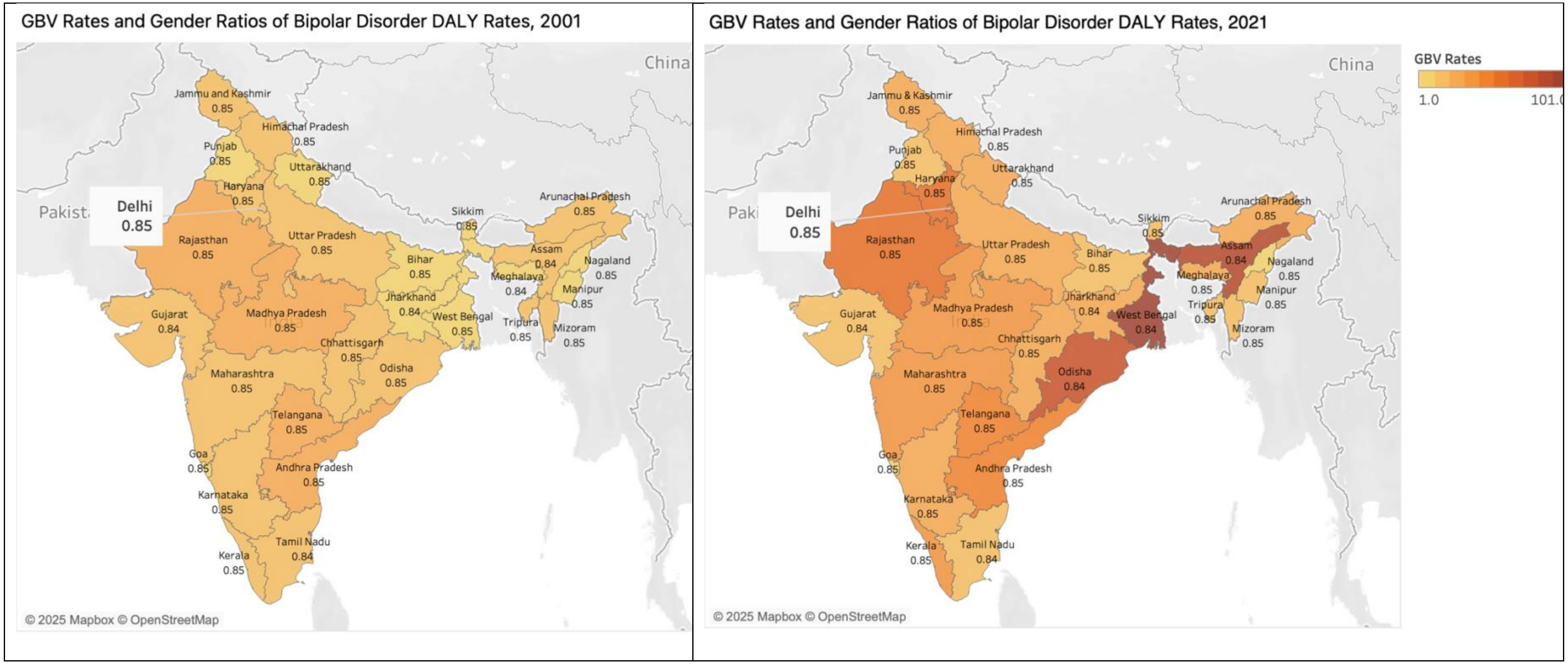

Note: The Indian map displays gender-based violence rates by colour gradient, with darker shades denoting higher rates and lighter shades denoting lower rates. Each geographical unit is labelled with the gender disparity ratio of bipolar disorder burden at baseline (2001) and the most recent year (2021). A gender disparity ratio greater than 1 denotes a higher bipolar disorder burden among women than men per 100,000 population.

**Figure S2.4 Distribution of gender-based violence rates and gender disparity in self-harm burden across 30 geographical units (Indian states/ Union Territories) at two time points, 2001 and 2021.**

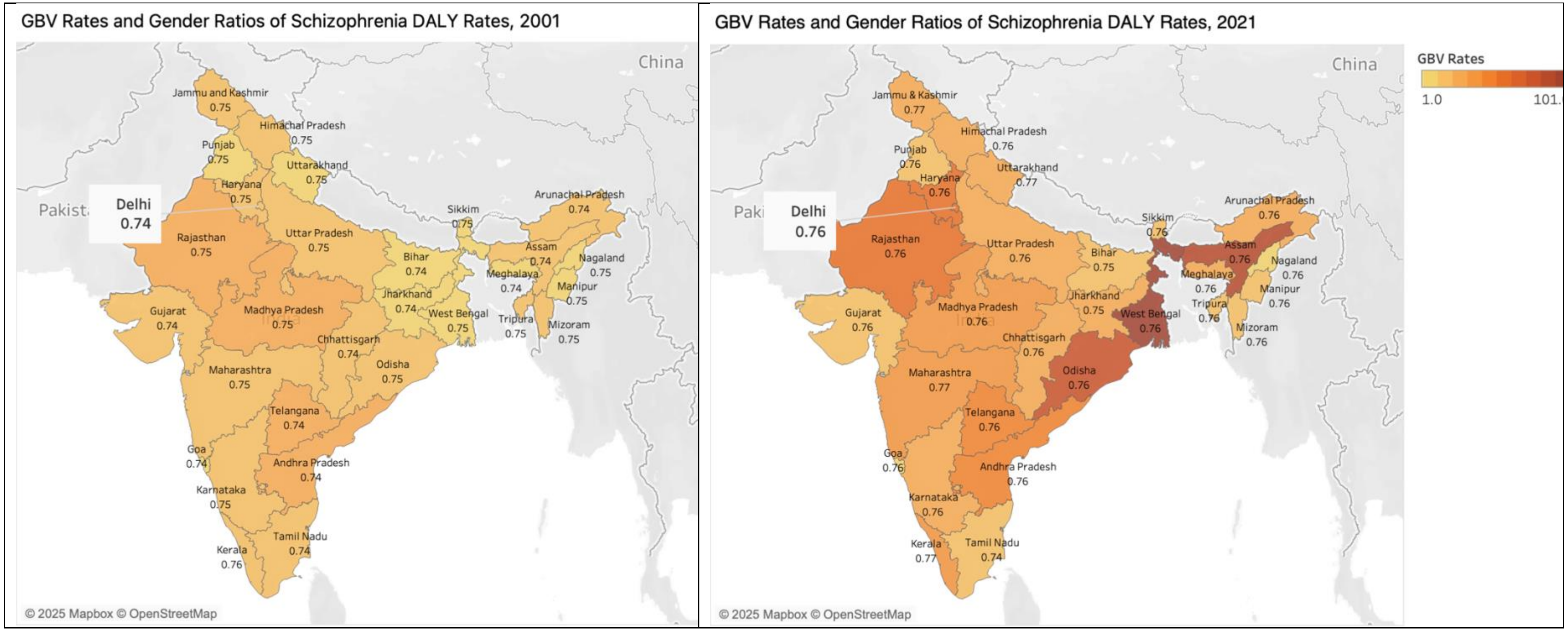

Note: The Indian map displays gender-based violence rates by colour gradient, with darker shades denoting higher rates and lighter shades denoting lower rates. Each geographical unit is labelled with the gender disparity ratio of schizophrenia burden at baseline (2001) and the most recent year (2021). A gender disparity ratio greater than 1 denotes a higher schizophrenia burden among women than men per 100,000 population.

**Figure S2.5 Distribution of gender-based violence rates and gender disparity in self-harm burden across 30 geographical units (Indian states/ Union Territories) at two time points, 2001 and 2021.**

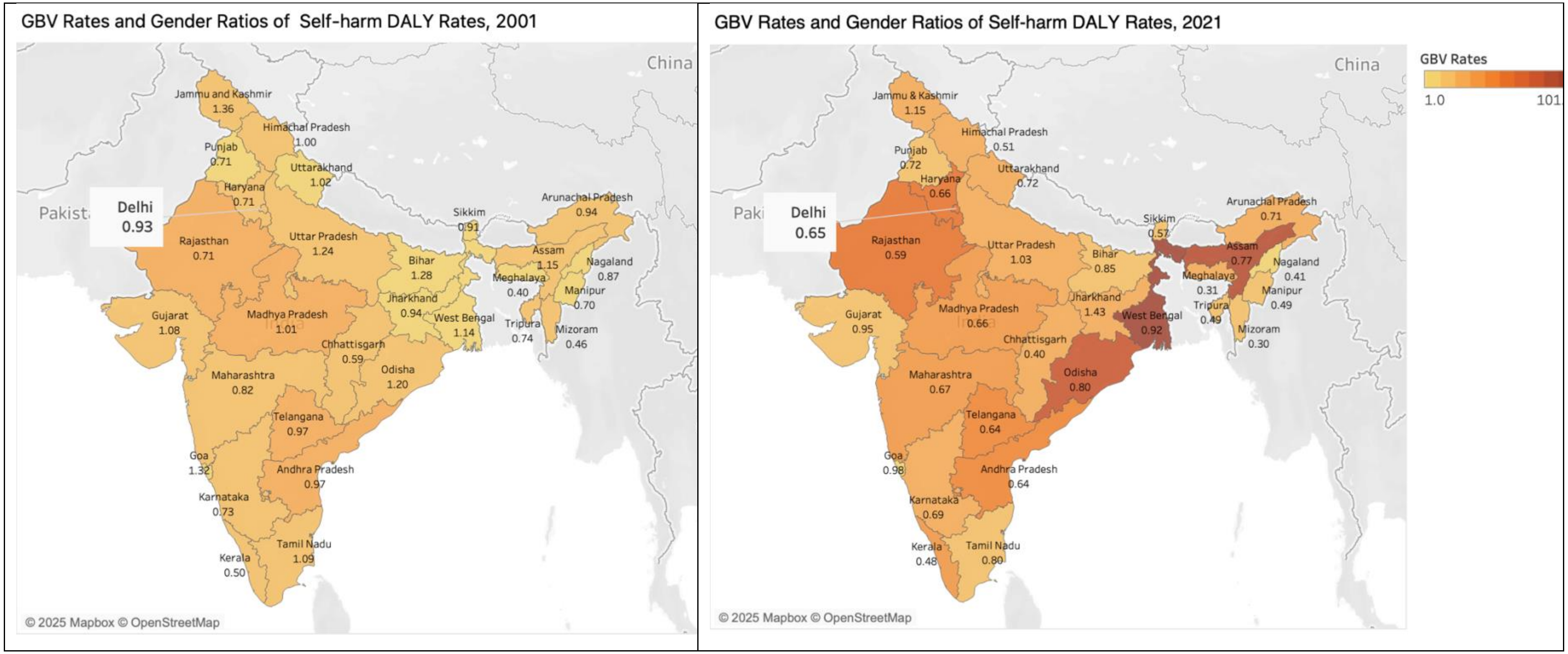

Note: The Indian map displays gender-based violence rates by colour gradient, with darker shades denoting higher rates and lighter shades denoting lower rates. Each geographical unit is labelled with the gender disparity ratio of self-harm burden at baseline (2001) and the most recent year (2021). A gender disparity ratio greater than 1 denotes a higher self-harm burden among women than men per 100,000 population.

**Figure S3.1 Line graph showing trends in gender-based violence rates and age-standardised disability-adjusted life year rates of schizophrenia across geographical units (Indian states/Union Territories) from 2001 to 2021.**

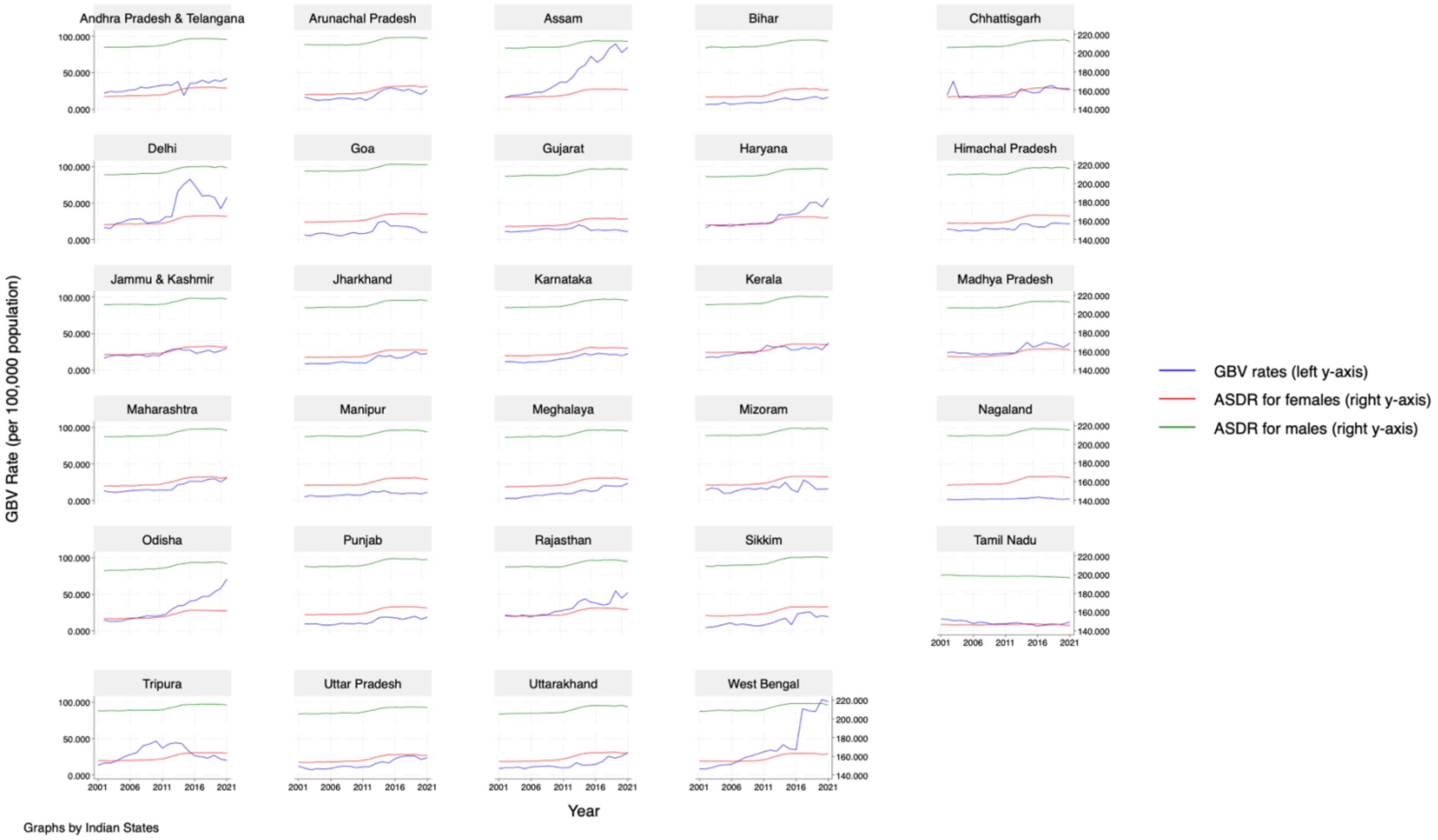

Note: Each panel shows one of the 30 geographical units (Indian states/Union territories). Andhra Pradesh and Telangana are combined in one panel. The x-axis denotes year from 2001to 2021. The left y-axis represents gender-based violence (GBV) rates per 100,000 population, plotted as blue lines. The right y-axis represents age-standardised disability-adjusted life year rates (ASDR) of schizophrenia for women/females (red lines) and men/males (green lines) per 100,000 population. Each panel illustrates state-specific changes in GBV rates and schizophrenia ASDRs over time from 2001 to 2021.

**Figure S3.2 Line graph showing trends in gender-based violence rates and age-standardised disability-adjusted life year rates of depressive disorders across geographical units (Indian states/Union Territories) from 2001 to 2021.**

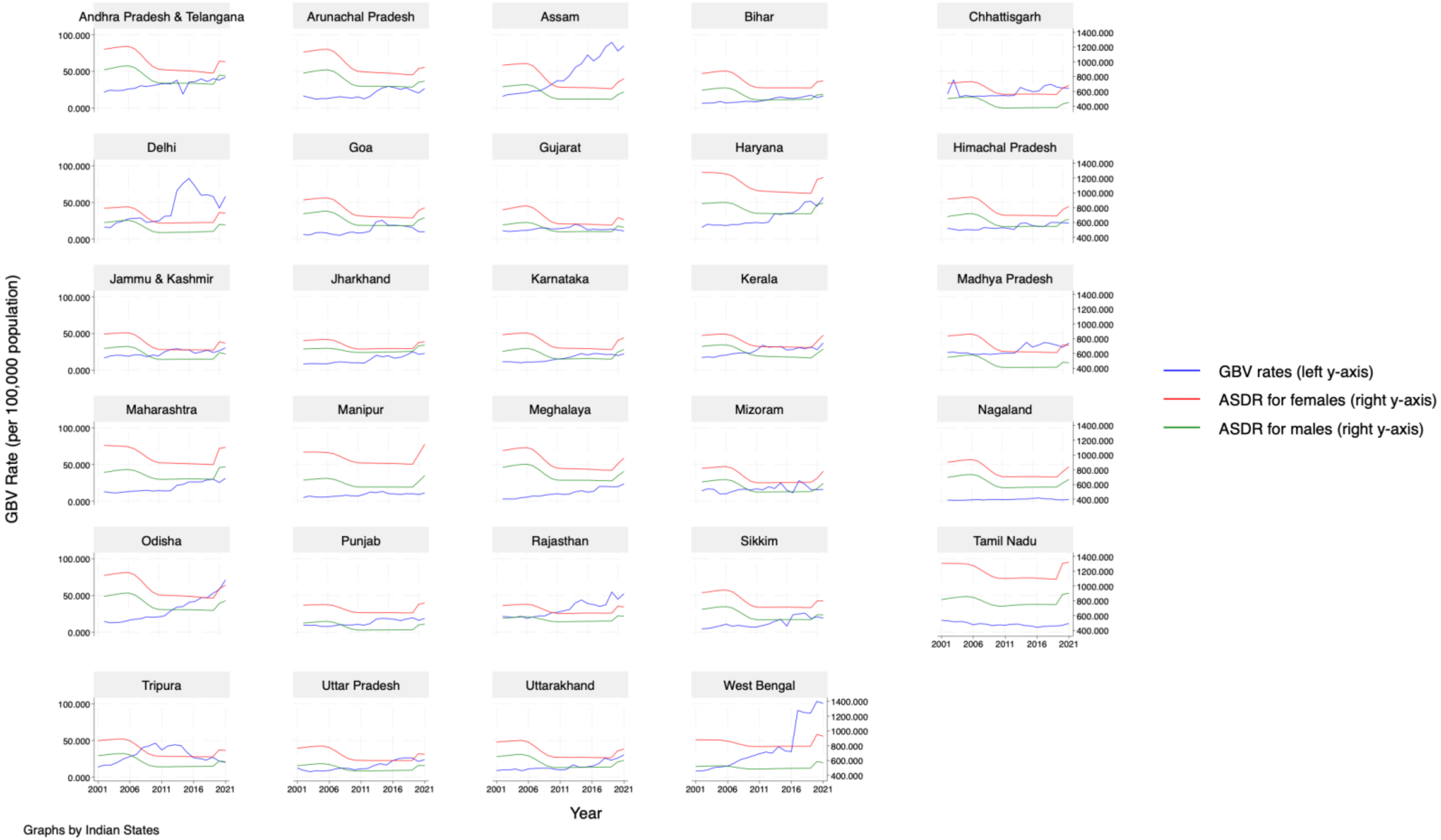

Note: Each panel shows one of the 30 geographical units (Indian states/Union territories). Andhra Pradesh and Telangana are combined in one panel. The x-axis denotes year from 2001to 2021. The left y-axis represents gender-based violence (GBV) rates per 100,000 population, plotted as blue lines. The right y-axis represents age-standardised disability-adjusted life year rates (ASDR) of depressive disorders for women/females (red lines) and men/males (green lines) per 100,000 population. Each panel illustrates state-specific changes in GBV rates and depressive disorder ASDRs over time from 2001 to 2021.

**Figure S3.3 Line graph showing trends in gender-based violence rates and age-standardised disability-adjusted life year rates of bipolar disorder across geographical units (Indian states/Union Territories) from 2001 to 2021.**

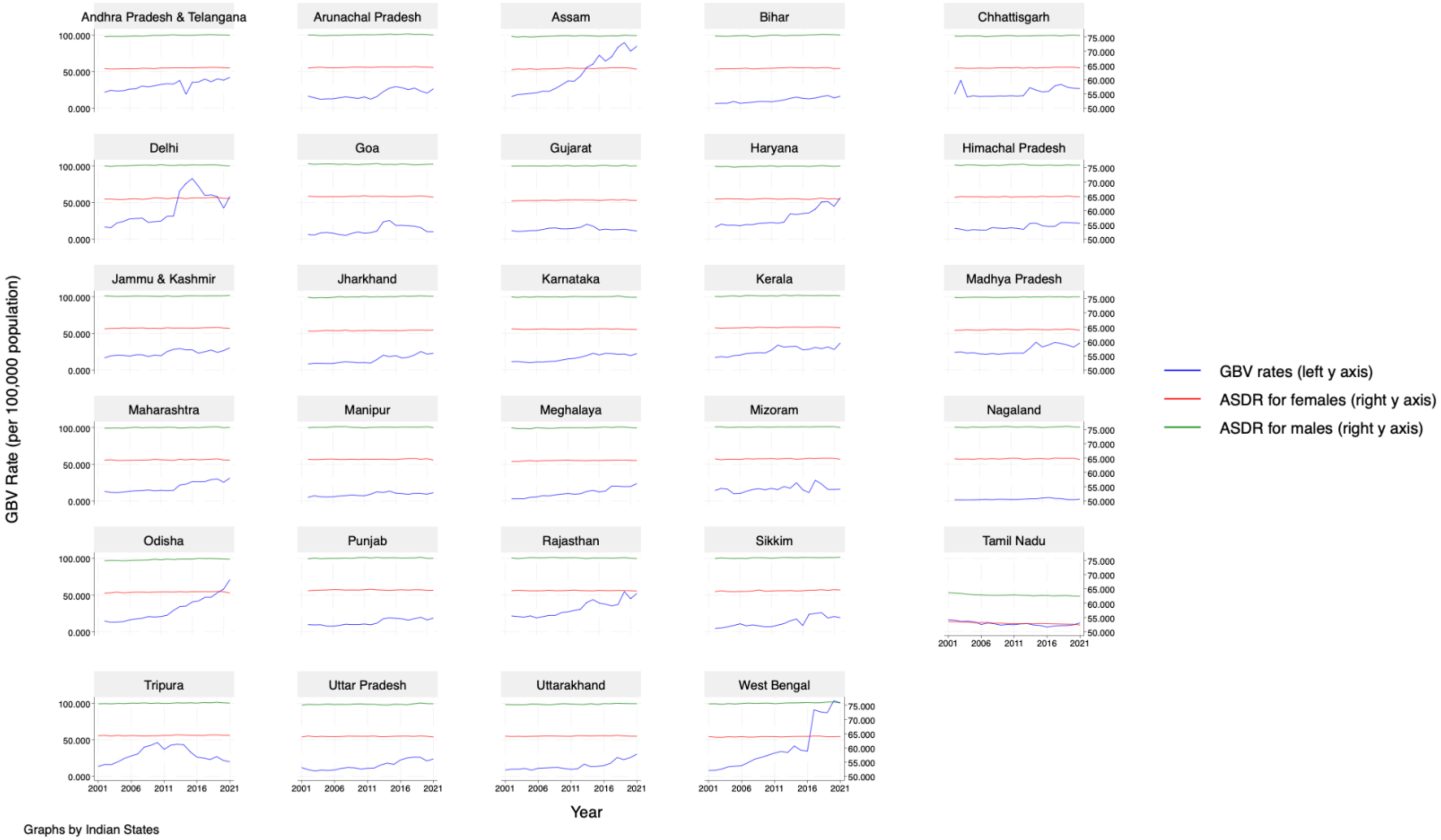

Note: Each panel shows one of the 30 geographical units (Indian states/Union territories). Andhra Pradesh and Telangana are combined in one panel. The x-axis denotes year from 2001to 2021. The left y-axis represents gender-based violence (GBV) rates per 100,000 population, plotted as blue lines. The right y-axis represents age-standardised disability-adjusted life year rates (ASDR) of bipolar disorder for women/females (red lines) and men/males (green lines) per 100,000 population. Each panel illustrates state-specific changes in GBV rates and bipolar disorder ASDRs over time from 2001 to 2021.

**Figure S3.4 Line graph showing trends in gender-based violence rates and age-standardised disability-adjusted life year rates of anxiety disorders across geographical units (Indian states/Union Territories) from 2001 to 2021.**

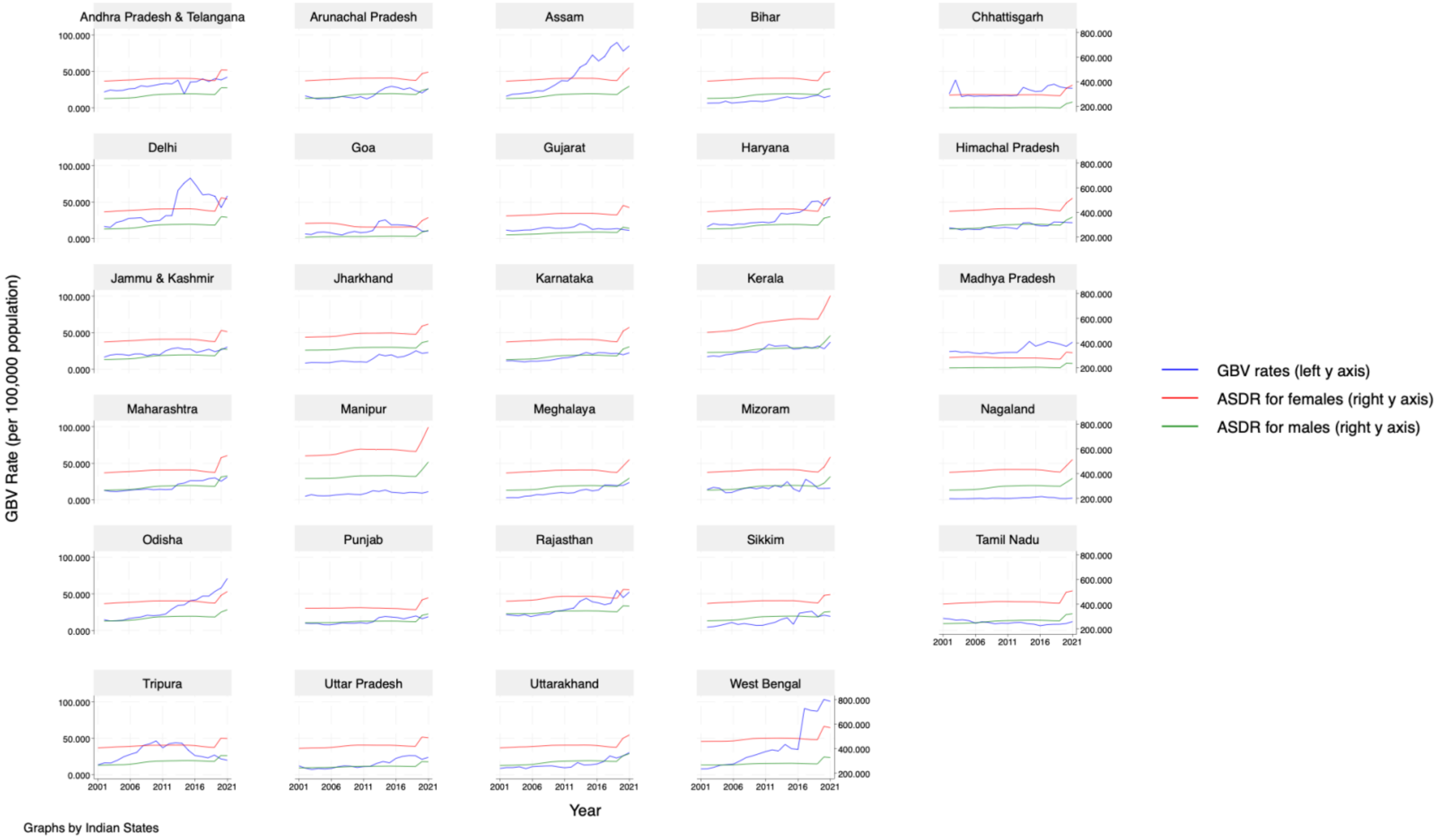

Note: Each panel shows one of the 30 geographical units (Indian states/Union territories). Andhra Pradesh and Telangana are combined in one panel. The x-axis denotes year from 2001to 2021. The left y-axis represents gender-based violence (GBV) rates per 100,000 population, plotted as blue lines. The right y-axis represents age-standardised disability-adjusted life year rates (ASDR) of anxiety disorders for women/females (red lines) and men/males (green lines) per 100,000 population. Each panel illustrates state-specific changes in GBV rates and anxiety disorder ASDRs over time from 2001 to 2021.

**Figure S3.5 Line graph showing trends in gender-based violence rates and age-standardised disability-adjusted life year rates of self-harm across geographical units (Indian states/Union Territories) from 2001 to 2021.**

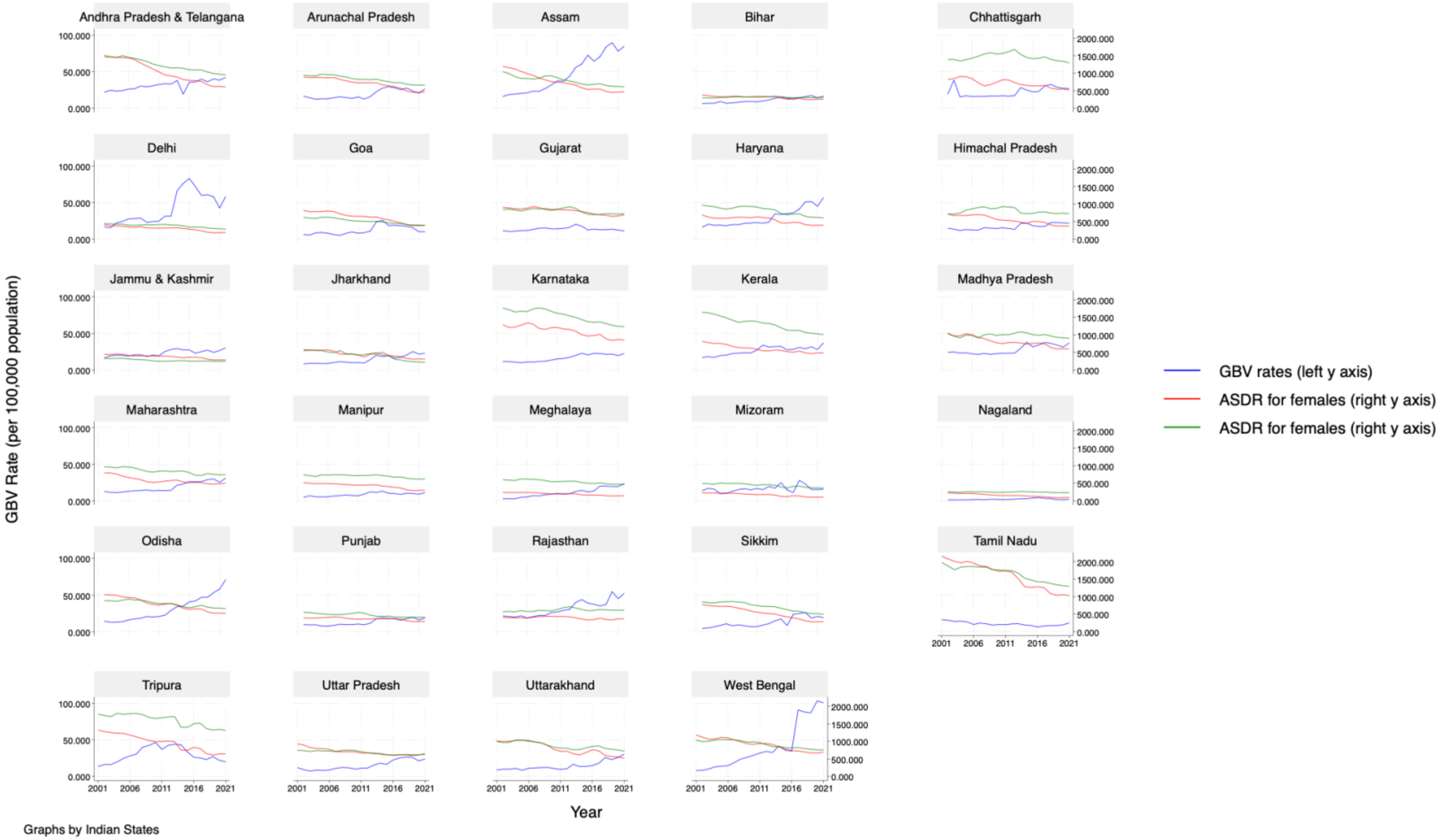

Note: Each panel shows one of the 30 geographical units (Indian states/Union territories). Andhra Pradesh and Telangana are combined in one panel. The x-axis denotes year from 2001to 2021. The left y-axis represents gender-based violence (GBV) rates per 100,000 population, plotted as blue lines. The right y-axis represents age-standardised disability-adjusted life year rates (ASDR) of self-harm for women/females (red lines) and men/males (green lines) per 100,000 population. Each panel illustrates state-specific changes in GBV rates and self-harm ASDRs over time from 2001 to 2021.

**4. Links for data input sources**

Link to metadata in Excel format can be found here.

| Year | Link to crime against women NCRB data from Open Government Data India portal | Link to Population estimates source |
|---|---|---|
| 2001 | https://www·data·gov·in/resource/stateut-city-wise-incidence-rate-crime-committed-against-women-during-2001 | n/a |
| 2002 | https://www·data·gov·in/resource/stateut-city-wise-incidence-rate-crime-committed-against-women-during-2002 | n/a |
| 2003 | https://www·data·gov·in/resource/stateut-city-wise-incidence-rate-crime-committed-against-women-during-2003 | n/a |
| 2004 | https://www·data·gov·in/resource/stateut-city-wise-incidence-rate-crime-committed-against-women-during-2004 | n/a |
| 2005 | https://www·data·gov·in/resource/stateut-city-wise-incidence-rate-crime-committed-against-women-during-2005 | n/a |
| 2006 | https://www·data·gov·in/resource/stateut-city-wise-incidence-rate-crime-committed-against-women-during-2006 | n/a |
| 2007 | https://www·data·gov·in/resource/stateut-city-wise-incidence-and-rate-crime-committed-against-women-during-2007 | n/a |
| 2008 | https://www·data·gov·in/resource/stateut-city-wise-incidence-rate-crime-committed-against-women-during-2008 | n/a |
| 2009 | https://www·data·gov·in/resource/stateut-city-wise-incidence-rate-crime-committed-against-women-during-2009 | n/a |
| 2010 | https://www·data·gov·in/resource/stateut-city-wise-incidence-rate-crime-committed-against-women-during-2010 | n/a |
| 2011 | https://www·data·gov·in/resource/stateut-wise-incidence-crimes-committed-against-women-during-2011 | n/a |
| 2012 | https://www·data·gov·in/resource/crime-against-women-during-2001-2012 | https://cbhidghs·mohfw·gov·in/WriteReadData/l892s/Demographic%20Indicators-2013·pdf |
| 2013 | https://www·data·gov·in/resource/crime-against-women-during-2013 | https://cbhidghs·mohfw·gov·in/WriteReadData/l892s/Demographic%20Indicators-2013·pdf |
| 2014 | https://www·data·gov·in/resource/crime-against-women-during-2014 | https://cbhidghs·mohfw·gov·in/WriteReadData/l892s/Demographic%20Indicators-2013·pdf |
| 2015 | https://www·data·gov·in/resource/state-ut-wise-cases-reported-i-victimsv-rate-r-crimes-committed-against-women-during-2015 | n/a |
| 2016 | https://www·data·gov·in/resource/stateut-wise-ipc-and-sll-crimes-against-women-during-2016 | n/a |
| 2017 | https://www·data·gov·in/resource/stateut-crime-head-wise-ipc-and-sll-crimes-against-women-during-2017 | n/a |
| 2018 | https://www·data·gov·in/resource/stateut-wise-crime-against-women-ipc-sll-2016-2018 | n/a |
| 2019 | https://www·data·gov·in/resource/stateut-and-crime-head-wose-ipc-crimes-against-women-during-2019 | n/a |
| 2020 | https://www·data·gov·in/resource/stateut-wise-crime-against-women-ipcsll-2018-2020 | n/a |
| 2021 | https://www·data·gov·in/resource/stateuts-wise-indian-penal-code-ipc-crimes-special-local-laws-sll-crimes-against-women | n/a |

**5. GATHER Checklist**

| Item | Checklist item | Reported on page # |
|---|---|---|
| **Objectives and funding** | | |
| 1 | Define the indicator(s), populations (including age, sex, and geographic entities), and time period(s) for which estimates were made. | Pg 4-5 |
| 2 | List the funding sources for the work. | n/a |
| **Data Inputs** | | |
| *For all data inputs from multiple sources that are synthesised as part of the study:* | | |
| 3 | Describe how the data were identified and how the data were accessed. | Pg. 4 |
| 4 | Specify the inclusion and exclusion criteria. Identify all ad-hoc exclusions. | n/a |
| 5 | Provide information on all included data sources and their main characteristics. For each data source used, report reference information or contact name/institution, population represented, data collection method, year(s) of data collection, sex and age range, diagnostic criteria or measurement method, and sample size, as relevant. | Pg.4-5 |
| 6 | Identify and describe any categories of input data that have potentially important biases (e.g., based on characteristics listed in item 5). | Pg. 8 |
| *For data inputs that contribute to the analysis but were not synthesised as part of the study:* | | |
| 7 | Describe and give sources for any other data inputs. | n/a |
| *For all data inputs:* | | |
| 8 | Provide all data inputs in a file format from which data can be efficiently extracted (e.g., a spreadsheet rather than a PDF), including all relevant meta-data listed in item 5. For any data inputs that cannot be shared because of ethical or legal reasons, such as third-party ownership, provide a contact name or the name of the institution that retains the right to the data. | supplement Pg. 25 |
| **Data analysis** | | |
| 9 | Provide a conceptual overview of the data analysis method. A diagram may be helpful. | - |
| 10 | Provide a detailed description of all steps of the analysis, including mathematical formulae. This description should cover, as relevant, data cleaning, data pre-processing, data adjustments and weighting of data sources, and mathematical or statistical model(s). | Supplement pg.3 |
| 11 | Describe how candidate models were evaluated and how the final model(s) were selected. | n/a |
| 12 | Provide the results of an evaluation of model performance, if done, as well as the results of any relevant sensitivity analysis. | Supplement pg. 10-11 |
| 13 | Describe methods for calculating uncertainty of the estimates. State which sources of uncertainty were, and were not, accounted for in the uncertainty analysis. | Pg. 5 |
| 14 | State how analytic or statistical source code used to generate estimates can be accessed. | - |
| **Results and Discussion** | | |
| 15 | Provide published estimates in a file format from which data can be efficiently extracted. | Appendix A |
| 16 | Report a quantitative measure of the uncertainty of the estimates (e.g. uncertainty intervals). | Pg. 6-7 |

| | | |
|---|---|---|
| **17** | Interpret results in light of existing evidence. If updating a previous set of estimates, describe the reasons for changes in estimates. | Pg. 8-9 |
| **18** | Discuss limitations of the estimates. Include a discussion of any modelling assumptions or data limitations that affect interpretation of the estimates. | Pg. 8-9 |

Source: Stevens GA, Alkema L, Black RE, Boerma JT, Collins GS, Ezzati M, et al.
Guidelines for accurate and transparent health estimates reporting: The GATHER statement.
Lancet. 2016. doi: http://dx.doi.org/10.1016/S0140-6736(16)30388-9
